# Genetics of the human microglia regulome refines Alzheimer’s disease risk loci

**DOI:** 10.1101/2021.10.17.21264910

**Authors:** Roman Kosoy, John F. Fullard, Biao Zeng, Jaroslav Bendl, Pengfei Dong, Samir Rahman, Steven P. Kleopoulos, Zhiping Shao, Jack Humphrey, Katia de Paiva Lopes, Alexander W. Charney, Brian. H. Kopell, Towfique Raj, David Bennett, Christopher P. Kellner, Vahram Haroutunian, Gabriel E. Hoffman, Panos Roussos

**Affiliations:** Pamela Sklar Division of Psychiatric Genomics, Icahn School of Medicine at Mount Sinai, NY, USA; Department of Genetics and Genomics Sciences, Icahn School of Medicine at Mount Sinai, NY, USA; Icahn Institute for Data Science and Genomic Technology, Icahn School of Medicine at Mount Sinai, NY, USA; Friedman Brain Institute, Icahn School of Medicine at Mount Sinai, NY, USA; Department of Psychiatry, Icahn School of Medicine at Mount Sinai, NY, USA; Department of Neurosurgery, Icahn School of Medicine at Mount Sinai, NY, USA; Department of Neuroscience, Icahn School of Medicine at Mount Sinai, NY, USA; Department of Neurology, Icahn School of Medicine at Mount Sinai, NY, USA; Rush Alzheimer’s Disease Center, Rush University Medical Center, Chicago, Illinois, USA; Department of Neurological Sciences, Rush University Medical Center, Chicago, Illinois, USA; Mental Illness Research Education, and Clinical Center (VISN 2 South), James J. Peters VA Medical Center, Bronx, NY, USA

## Abstract

Microglia are brain resident myeloid cells that play a critical role in neuroimmunity and the etiology of Alzheimer’s Disease (AD). Yet our understanding of how the genetic regulatory landscape controls microglial function and contributes to disease is limited. Here, we performed transcriptome and chromatin accessibility profiling in primary human microglia from 150 donors to identify genetically-driven variation and cell-specific enhancer-promoter interactions. Integrative fine-mapping analysis identified putative regulatory mechanisms for 21 AD risk loci, of which 18 were refined to a single gene, including 3 novel genes (*KCNN4, FIBP* and *LRRC25*). Transcription factor regulatory networks captured AD risk variation and identified *SPI1* as a key regulator of microglia expression and AD risk. This comprehensive resource capturing variation in the human microglia regulome provides novel insights into the etiology of neurodegenerative disease.

**One-Sentence Summary:** Characterizing the genetic regulation of chromatin accessibility and gene expression in human microglia refines molecular mechanisms of Alzheimer’s disease risk loci.

---

Microglia are resident macrophage-like cells constituting ∼5–10% of all brain cells. Microglia display a diverse range of functions, mediated through interactions with neighboring glial and neuronal cells ^1^. There is an increasing focus on understanding the molecular and genetic mechanisms involved in microglia function as they are central to multiple neurodegenerative disorders, including Alzheimer’s disease (AD), Parkinson’s disease (PD), multiple sclerosis (MS), and amyotrophic lateral sclerosis (ALS) ^2,3^. However, studying the regulatory and transcriptional mechanisms of human primary microglia is challenging as fresh brain material is not readily available.

Previous efforts have established the importance of microglia regulatory elements in the etiology of AD due to the enrichment of AD risk variants within regions of microglia specific accessible chromatin ^4–6^. Expression quantitative trait loci (eQTL) datasets from primary microglia ^7,8^ can help to map functional AD risk variants and nominate target genes. Genetic analysis of variation in chromatin accessibility will significantly enhance these efforts by identifying AD risk variants that directly affect transcriptional cis-regulatory activity, revealing the microglia-specific regulatory mechanisms disrupted in disease.

In the current study, we performed population-based analysis of the human microglia regulome and transcriptome to understand the role of transcription factor (TF) regulatory networks and the genetic regulatory landscape implicated in neurodegenerative diseases. We generated multi-omics data in microglia isolated from fresh human brain tissue of 150 unique donors and used this to develop an atlas of chromatin accessibility and to examine microglia-specific enhancer-promoter interactions. We then examined the population-level variation of gene expression and chromatin accessibility, and jointly utilized these resources to investigate the genetically driven regulation of transcription in microglia. This approach enabled us to fine-map AD loci, identify novel putative AD risk genes and provide mechanisms for how disease-associated risk variants contribute to the dysregulation of expression of microglia genes relevant to AD.

## Landscape of chromatin accessibility and gene expression in primary human microglia

We performed genotyping and generated ATAC-seq (n=107), RNA-seq (n=127) and Hi-C (n=5) in primary human microglia isolated from fresh prefrontal cortex tissue from a total of 150 unique donors derived from biopsies (n=27) and autopsies (n=123) (**Fig. 1a, Fig S1, Table S1**,). Microglia were isolated by fluorescent activated cell sorting (FACS) of viable CD45^+^ and CD11b^+^ positive cells from dissociated brain specimens (**Fig S2, see Methods**). After data preprocessing, we retained 210,832 open chromatin regions (OCRs) (**Fig. S3a**) and 18,856 genes for further analyses, with 88 samples having high quality data for both ATAC-seq and RNA-seq (**Fig S4)**. Our ATAC-seq (**Fig. 1b**) and RNA-seq (**Fig. S5**) data clustered closely with microglia from previous studies ^4,9,10^ (**Table S2**).

**Fig. 1.**
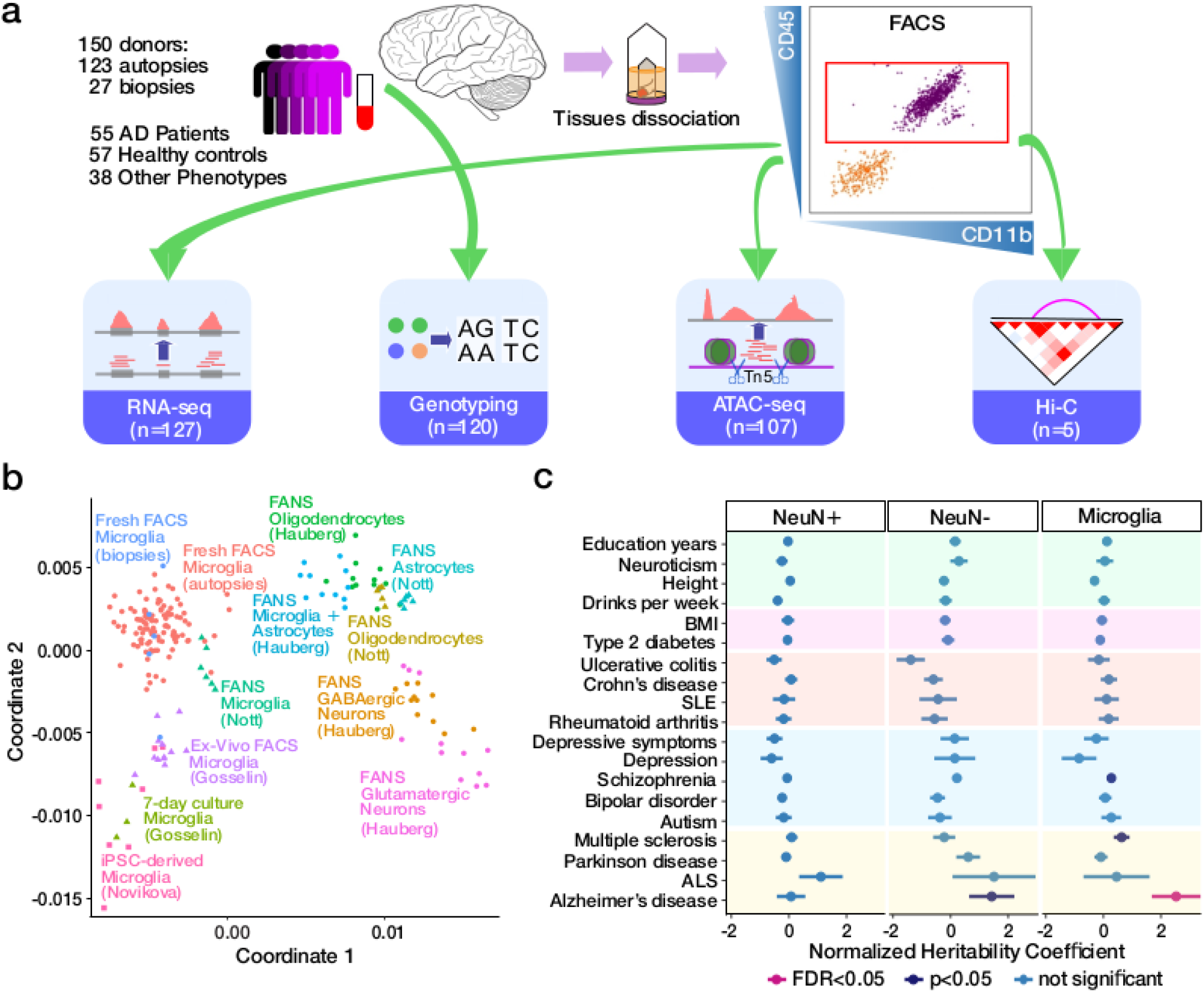
Chromatin accessibility landscape in human microglia and AD predisposition. **a)** Schematic outline of data generation. **b)** Comparison of human microglia ATAC-seq dataset to other brain open chromatin datasets (**Table S2**) utilizing jointly called OCRs in multidimensional scaling space. **c)** Enrichment of trait-associated genetic variants in neuronal (NeuN+), non-neuronal (NeuN-), microglia and microglia-specific OCRs. Coefficients from LD score regression are normalized by the per-SNP heritability (h2 / total SNPs per GWAS). Horizontal bars indicate standard error.

We examined the overlap of our population-scale chromatin accessibility map with existing OCR datasets and genetic risk variants. Chromatin accessible regions from our data had higher overlap with microglia-specific OCRs (Jaccard *J*=0.366) relative to those from other brain cell populations (Jaccard *J* between 0.138-0.178) ^6^ (**Fig. S3b**). While observed enrichment was highest for promoters, distal OCRs showed the highest specificity for microglia and is consistent with higher cell type-specificity associated with distal regulation ^11^. The relevance of microglia OCRs to human diseases was evaluated by examining enrichment for common genetic risk variants. Consistent with previous studies ^4,6,9,12^, we observed an enrichment of AD risk variants ^12^ specifically in microglia OCRs (**Fig. 1c**) (**Table S3**). Furthermore, microglial OCRs explained higher AD heritability (FoldChange=4.0, p=0.013, one-sided two-sample z-test) compared to OCRs discovered in broadly defined populations of non-neuronal cells^5^.

## Transcriptional regulation by open chromatin regions

We next evaluated the coordination between the genome-wide OCR landscape and transcriptional activity in microglia using our unique resource of chromatin accessibility and gene expression data from the shared set of 88 donors. We fit a variance decomposition model for each gene to estimate the fraction of expression variation attributable to genome-wide variation in chromatin accessibility. Analysis of 185,664 OCRs, located within a 100kb window centered around transcription start sites (TSS), revealed that variation in chromatin accessibility explained a median of 83.4% of expression variation across 18,640 genes, compared to a median of 0% explained in permuted data (p<10^−323^, one-sided Wilcoxon test) (**Fig. 2a**). Variation in chromatin accessibility explained at least 75% of transcriptional variance for 83.1% of the investigated genes (15,491), indicating strong coupling between chromatin structure and gene expression in human microglia (**Fig. S6**).

**Fig. 2.**
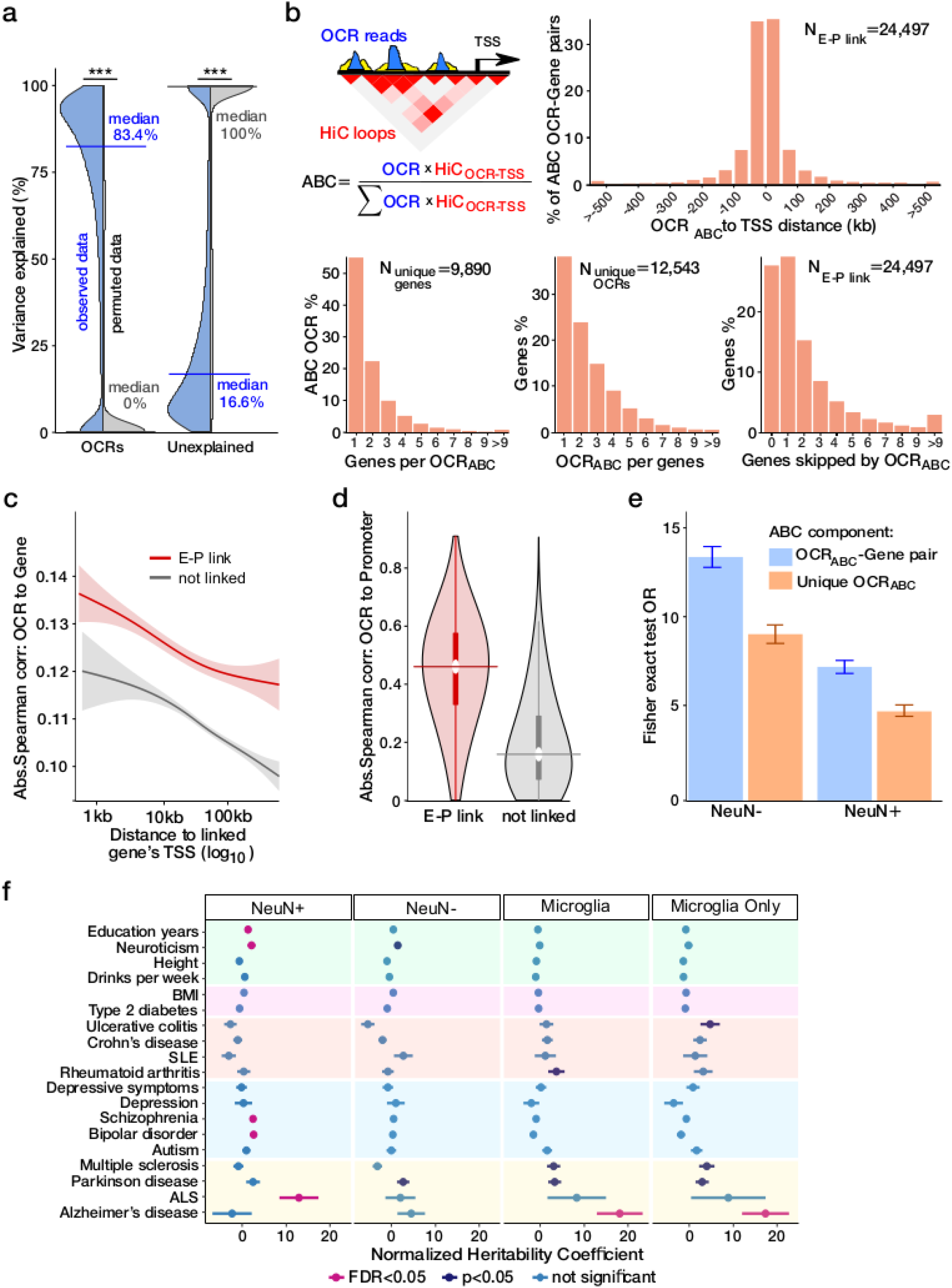
Transcriptional regulation by open chromatin regions. **a)** Fraction of transcriptional variation for each gene explained by accessible chromatin for observed data (blue) and permuted data (grey). **b)** Distribution of distance from TSS for E-P interactions (top right); histograms of the number of OCR_ABC_ per gene (bottom left), the number of genes per OCR_ABC_ (bottom middle) and the number of skipped genes between the OCR_ABC_ and the linked gene (bottom right). **c)** OCR_ABC_ involved in E-P interactions have stronger correlation with the expression of the corresponding gene compared to non E-P pairs. **d)** OCR_ABC_ involved in E-P interactions have stronger correlation with the OCR at the linked promoter compared to OCRs not in an E-P link. Horizontal lines indicate the median, and thick vertical lines indicate 25%-75% quantiles. **e)** Enrichment of microglia E-P interactions with non-neuronal (NeuN-) and neuronal (NeuN+) E-P interactions. Colored bars indicate the odds ratio and error bars represent 95% confidence intervals. **f)** Enrichment of trait-associated genetic variants in neuronal (NeuN+), non-neuronal (NeuN-), microglia and microglia-specific E-P interactions. Coefficients from LD score regression are normalized by the per-SNP heritability (h2 / total SNPs per GWAS). Horizontal bars indicate standard error.

Next, with the aim of linking a regulatory element to the gene(s) it regulates, we used a recently developed “activity-by-contact” (ABC) framework ^13^ to combine our Hi-C derived contact frequencies with “enhancer activities” in OCRs to examine long-range enhancer-promoter (E-P) interactions (**Fig. 2b**). We identified 24,497 E-P interactions, involving 9,890 unique genes, thus identifying at least one non-promoter regulatory element for over 52% of microglia expressed genes. About half of the E-P linked OCRs, termed OCR_ABC_, were linked to more than one gene, and over 60% of the linked genes were linked to multiple OCR_ABC_ (**Fig. 2b**). As demonstrated previously (*15*), OCR_ABC_ more often did not target the nearest gene (72% OCR_ABC_ skipped at least one gene) (**Fig. 2b**), further highlighting the importance of experimentally derived regulatory annotations.

OCR_ABC_ have a significantly higher correlation with the expression of linked genes compared to unlinked genes (p<10^−95^, one-sided Wilcoxon test) (**Fig. 2c**, as well as to chromatin accessibility at the linked promoter compared to OCRs that do not participate in E-P interactions (p<10^−323^, one-sided Wilcoxon test) (**Fig. 2d**). As expected ^13^, the majority of observed E-P interactions corresponded to a positive correlation between gene expression and chromatin accessibility (**Fig. S7a-b**). To evaluate the cell type specificity of the E-P interactions observed in microglia, we compared them to E-P pairs identified in broad neuronal (38,233 pairs) and non-neuronal (37,056 pairs) cell populations ^14^. In total, 23.6% (5,781 out of 24,459) microglia E-P interactions were shared with either neurons or non-neurons (**Fig. S8a-b**). As expected, we observed a stronger overlap between microglial and non-neuronal (OR=13.7) E-P interactions than with those of neurons (OR=7.5) (**Fig. 2e; Fig. S8c**). Conversely, over 76% of the E-P interactions were observed in microglia alone, reflecting the cell-type specificity of regulatory mechanisms.

To further explore the importance of cell type-specific regulatory mechanisms and their role in disease, we quantified the overlap of disease risk variants with OCR_ABC_ from neurons, glia, and microglia. Similar to analysis of all OCRs above (**Fig. 1f**), enrichment of AD risk variants was only observed in microglia OCR_ABC_ (**Fig. 2f**). Strikingly, limiting analysis to microglial E-P interactions increases the explained heritability coefficient for AD (fold change=7.2, p=0.0016, one-sided two-sample z-test). This highlights the central role for transcriptional regulatory mechanisms of human microglia in the genetic architecture of AD.

## Genetic regulation of chromatin accessibility in human microglia

ATAC-seq and high-density genotyping data from 95 donors allowed us to study population variation by generating a human microglia chromatin accessibility QTL (caQTL) map. Utilizing the multivariate multiple QTL (mmQTL) method ^15^, and correcting for multiple technical confounds (**Fig. S9)** and population structure, we used a 50kb window centered on each of 210,832 OCRs and identified 5,468 OCRs with significant caQTLs. Our microglia caQTL dataset had high concordance with caQTLs identified in human iPSC-derived macrophages, derived under various stimulating conditions (range of π1 values: 0.662 to 0.753), reflecting the shared myeloid origin of microglia and macrophages ^16^ (**Fig. S10a**). The replication of microglia caQTLs was lower in caQTLs derived from homogenate brain specimens (π_1_=0.602) ^17^.

Given the high concordance of caQTLs among microglia and myeloid cells, we maximized statistical power for caQTL detection by jointly analyzing our human microglia and four macrophage datasets ^16^. The resulting human microglia meta-caQTL dataset contained 10,266 OCRs with significant caQTLs. Bayesian analysis of results from these two cell types indicate that the majority of caQTLs were either discovered in microglia alone or had comparable level of support from the macrophage subsets ^18^ (**Fig. 3a**). By applying a fine-mapping approach ^19^ to the meta-caQTL results, we identified a 95% credible set of 269,536 SNPs, including 144,592 SNPs (called caSNPs) with posterior probability (PP) >0.01 for 10,152 OCRs (**Fig. S11a**). Of these, 6,476 caSNPs were located within 4,324 OCR peaks (**Fig. S12a-b**).

**Fig. 3.**
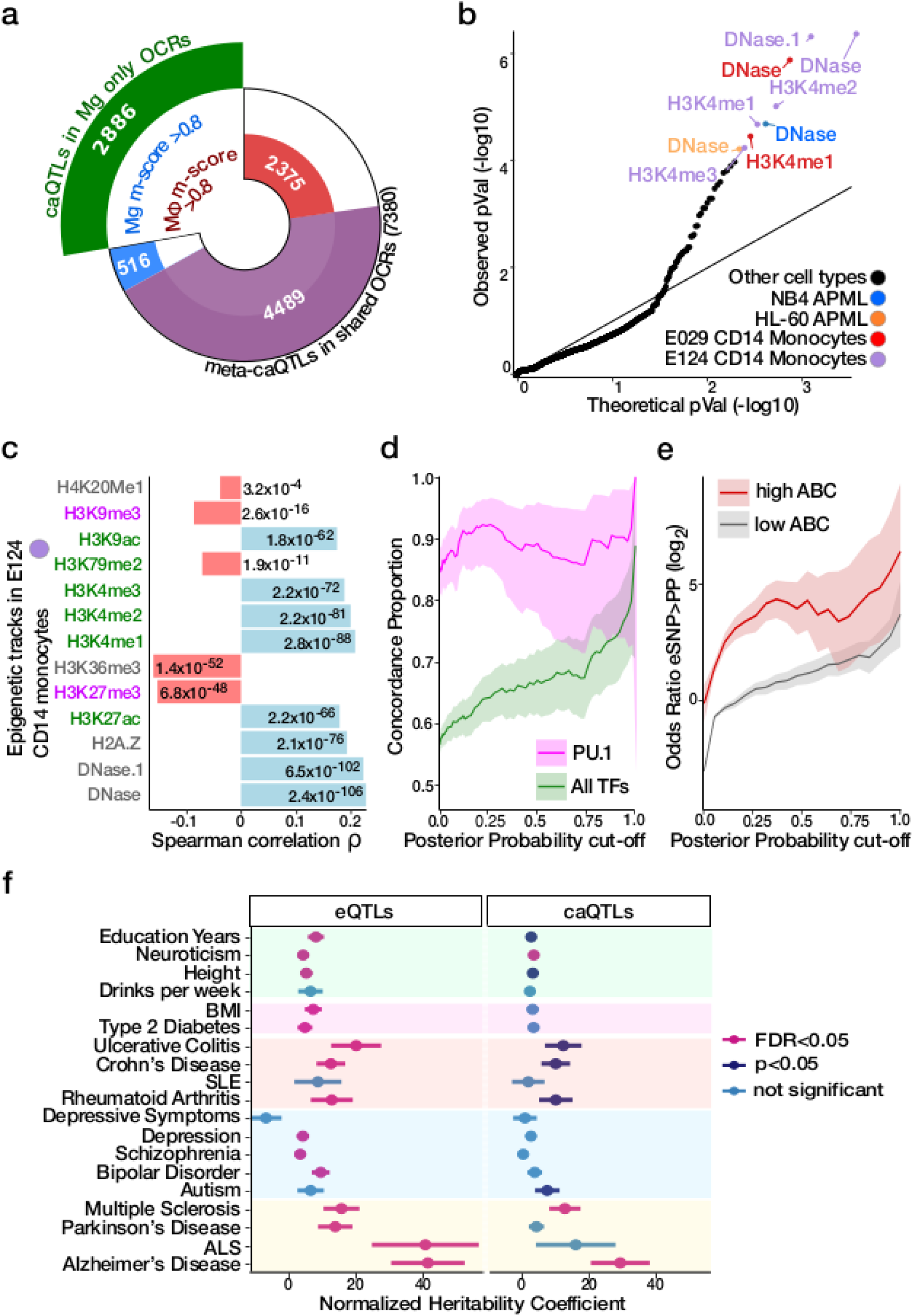
Genetic regulation of chromatin accessibility in human microglia. **a)** Count of OCRs with caQTL signals in microglia (Mg) and macrophages (Mφ) shown by cell type specificity based on Bayesian meta-analysis. Analysis of microglia-only OCRs gives caQTLs specific to microglia (green), and analysis of shared OCRs gives both shared and cell type specific caQTLs. **b)** QQ plot of p-values reflecting the concordance between DeepSEA predictions and caQTL regression coefficient. Significant assays from myeloid lineages are indicated by colors. **c)** Spearman correlation between caSNPs’ effect size estimated by caQTL analysis and by DeepSEA predicted effect on epigenetic assays for promoters/enhancers (green) and repressors (purple). P-values for each test are indicated. **d)** Concordance between caSNPs’ allelic effects on chromatin accessibility and the predicted change in motif binding ability for PU.1 compared to all 53 TFs (including PU.1), whose binding sites were significantly disrupted by caSNPs. Concordance is shown as a function of posterior probability from fine-mapping, and shaded regions indicate 95% confidence intervals. **e)** Enrichment for fine-mapped caSNPs within OCRABC also being fine-mapped eSNPs for the target genes compared to those in OCRs not involved in E-P interactions. Enrichments are shown over a range of posterior probability cutoffs applied to both caSNPs and eSNPs. Shading indicates 95% confidence interval. **f)** Enrichment of trait-associated genetic variants in 95% credible set of microglia meta-eSNPs and meta-caSNPs. Coefficients from LD score regression are normalized by the per-SNP heritability (h2 / total SNPs per GWAS). Horizontal bars indicate standard error.

Since genetic regulatory architecture varies across tissues and cell types ^20^, we evaluated the cell type specificity of the fine-mapped caSNPs with PP>0.01 and, within OCRs, by querying the predicted effect of each variant on 2,002 epigenetic assays across tissues and cell lines from ENCODE and the Roadmap Epigenomics Project estimated by DeepSEA ^21,22^. Even without considering any prior knowledge about the cell type of origin, the epigenetic tracks predicted to be most disrupted by this set of caSNPs were DNAse hypersensitivity sites (DHS) from primary CD14^+^ monocytes (**Fig. 3b**). Assayed epigenetic tracks in other myeloid lineage cell types were also disrupted. Moreover, the direction of the predicted effect was consistent with the known biology of these assays. For example, caQTL and DeepSEA effect directions were positively correlated with changes in DHS and ChIP-seq marks indicative of promoter and enhancer activity from myeloid lineages, and negatively correlated with changes in repressive epigenetic marks (**Fig. 3c**).

We evaluated the degree to which genetic variants affecting chromatin accessibility acted by disrupting transcription factor binding sites (TFBS). We employed a TF footprinting approach ^23^ to identify all bound TFs within the microglia accessible chromatin landscape. caSNPs were more likely to be located within occupied TFBSs, as determined by footprinting analysis (OR=1.10, p=6.95×10^−5^, Fisher’s exact test). We identified 53 TFs whose predicted binding was significantly enriched for dysregulation by caSNPs, as compared to non-caSNP genetic variants present within OCRs (**Table S4**). Among these, predicted SPI1 binding sites were the most significantly disrupted by caSNPs (OR=5.56, FDR=5.65×10^−55^, Fisher’s exact test) and the effect direction from caQTL analysis was concordant with the predicted TFBS disrupting allele at 92% of caSNPs (p=6.6×10^−4^, binomial test) (**Fig. S13**). While increasing the fine-mapping posterior probability cutoff for caSNPs increased the concordance for all TFs, predicted *SPI1* binding sites remained remarkably concordant across a wide range of cutoffs (**Fig. 3d**). The *SPI1* gene encodes PU.1, which is a master regulator of myeloid cell development and critical for microglia function ^24^. Our observation reinforces the importance of genetic regulation of *PU*.*1* target genes in human primary microglia.

## Coordinated genetic regulation of chromatin accessibility and gene expression

Using the same approach as for caQTLs, eQTL analysis on samples from 101 donors identified 1,603 eQTLs at 5% FDR (**Fig. S9b**). These microglia eQTLs were replicated in two other human microglia eQTL datasets (range of π_1_: 0.62 to 0.70) ^8,25^ (**Fig. S10b**). Given the high concordance, we performed meta-analysis and statistical fine-mapping of these 3 datasets, and identified 7,302 meta-eQTLs (**Fig. S9b; Fig. S12c**). Fine-mapped eSNPs were overrepresented within OCR_ABC_ corresponding to their target genes (OR=1.48, permutation test p=6.7×10-5) (**Fig. S14**).

Having shown that eSNPs are more likely to colocalize with E-P interaction, we next sought to examine the genetically driven regulation of transcription in microglia. Fine-mapped caSNPs within OCR_ABC_ regions were more likely to be fine-mapped eSNPs for the target gene compared to caSNPs for OCRs not involved in E-P interactions (**Fig. 3e**). Genetic colocalization analysis ^26^ identified 1,457 instances where pairs of gene expression and chromatin accessibility traits shared genetic regulatory architecture with high posterior probability (i.e. PP4>0.5, including 865 unique genes and 1,033 unique OCRs (**Fig. S15a**). Of these, 167 OCR-gene pairs were also E-P links identified via ABC (**Fig. S15c**). OCRs predicted to be involved in E-P interactions were enriched for being colocalized with an eQTL for the target gene (OR=2.5, permutation test p=6.7×10^−5^) (**Fig. S15d**).

Taken together, we have captured variation of the regulatory mechanisms that are involved in microglia E-P interactions which we further integrated with risk loci across multiple traits. We evaluated the overlap of microglia regulatory variants in the 95% credible sets for gene expression and chromatin accessibility with risk variants for common diseases. While eSNPs showed significant enrichment for a range of neurodegenerative, inflammatory and neuropsychiatric traits (15 of 20 tested), caSNPs showed a more specific signal with three traits, including the largest heritability coefficient for AD (**Fig. 3f**). Integrating fine-mapping for chromatin accessibility and gene expression (**see Methods**) produced a refined set of 30,028 variants showing significant enrichment for AD risk loci (p=0.036) (**Fig. S16**), pointing to a key role for the genetic regulation of gene expression via chromatin accessibility in AD.

## Integration of microglia regulome with AD risk variation

Having shown the high specificity of the microglia regulome for AD genetic risk architecture, we performed fine-mapping to better identify AD credible causal variants, genes and regulatory regions. We first examined the colocalization of fine-mapped AD risk variants ^12^ within microglia E-P interactions. Remarkably, 6,428 distal OCR_ABC_ (>20kb from the nearest TSS) contained 20 fine-mapped SNPs (PP>0.01) from eight different AD loci, while the 97,513 of the equidistant OCRs with low ABC scores contained none of the fine-mapped SNPs (OR=318, p=8.9×10^−25^, Fisher’s exact text) (**Fig. 4a; Table S5**).

**Fig. 4.**
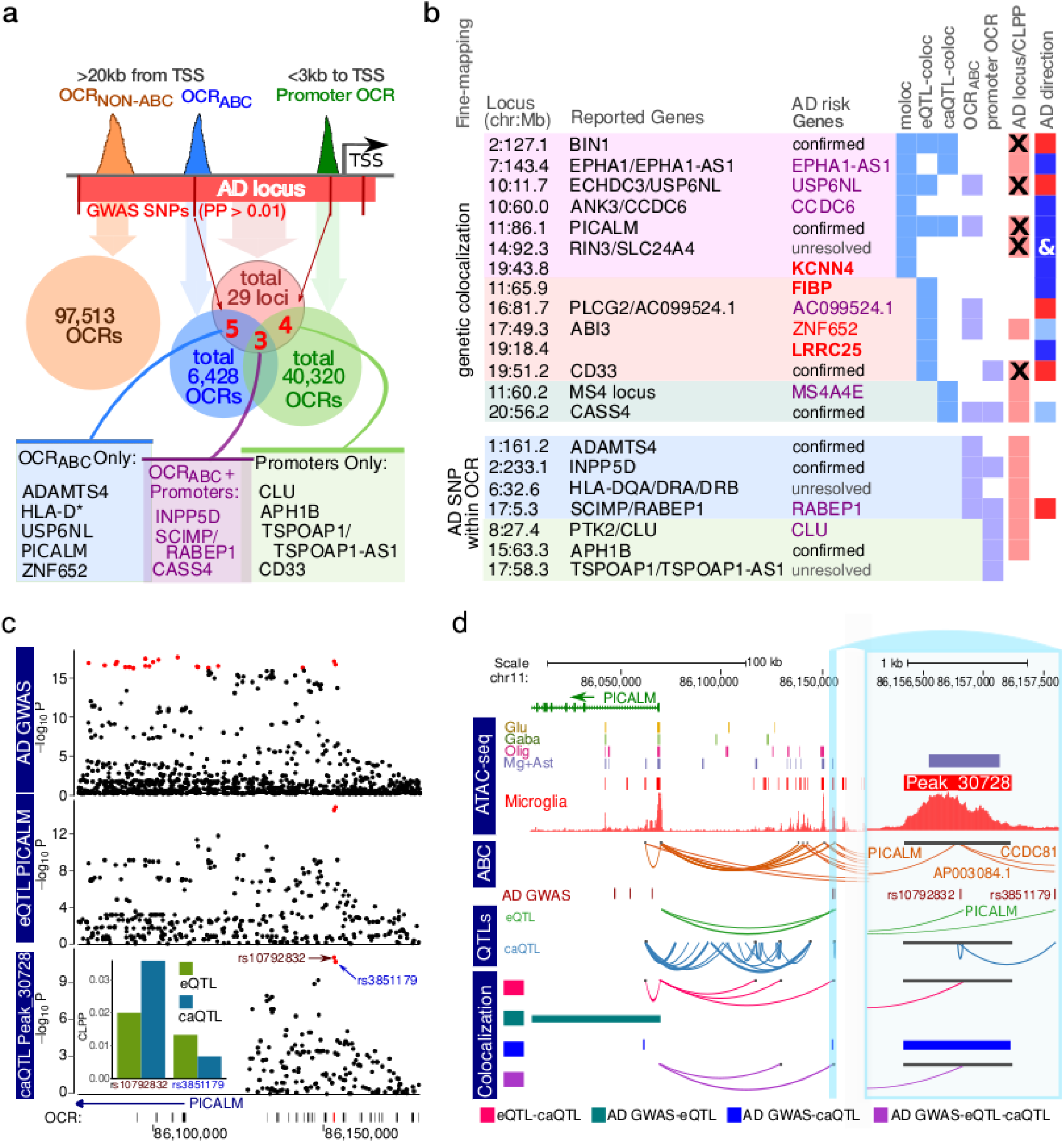
Integration of AD etiologic landscape with genetic regulation of transcriptional andomatin accessibility in microglia. **a)** Overlap of 316 fine-mapped SNPs from 29 AD GWAS loci ^12^ with OCRABC (blue) and promoters (green); **b)** Fine-mapping to define candidate AD genes based on: a) joint colocalization for eQTL, caQTL and GWAS signal (‘moloc’); colocalization for (b) eQTL and GWAS (‘eQTL-coloc’); and (c) caQTL and GWAS (‘caQTL-coloc’) signal; fine-mapped AD variants (PP>0.01) within (d) OCR_ABC_ and (e) promoter OCR. ‘AD GWAS’ indicates regions identified by Jansen *et al*. ^12^, and ‘x’ indicates significant joint fine-mapping with gene expression or chromatin accessibility. ‘AD direction’ is the linked gene’s expression in relation to the AD risk alleles (red=higher; blue=lower, ‘’ indicates consistency for multiple genes in the region). Color schema for ‘Linked Genes’: genes are unambiguously fine-mapped and previously implicated in AD (purple); not previously fine-mapped as AD risk genes (red). Novel putative AD risk genes outside previously reported AD loci are shown in bold. **c)** Local plot showing results from AD GWAS ^12^, eQTL analysis of PICALM, and caQTL analysis of peak_30728. Red points indicate genetic variants in the 95% credible set from statistical fine-mapping of each trait. Inset shows colocalization posterior probabilities (CLPP) for the top variants in the credible set for gene expression and chromatin accessibility. **d)** Visualization of the PICALM locus showing: open chromatin regions from 4 cell populations ^5^ and microglia from this study; E-P interactions (ABC); fine-mapped (PP>0.05) SNPs from AD GWAS ^12,39,44^; genetic regulation from eQTLs and caQTL form thus study; and colocalization analysis between pairs of traits (i.e. AD GWAS, gene expression chromatin accessibility) using ‘coloc’ and all three traits using ‘moloc’ methods.

We then combined AD genetic risk variation ^12^ with the microglia meta-eQTL and meta-caQTL datasets using multiple-trait-coloc (moloc) ^27^ to link AD loci to genes and regulatory regions (**Table S6)**. We observed GWAS-eQTL-caQTL colocalization within six previously reported AD loci (**Fig. 4b)**, providing coherent units of transcriptional regulation relevant to the etiology of AD. Colocalization analyses between the AD GWAS and meta-eQTL and meta-caQTLs, separately, provided functional annotation for five additional published AD loci. Importantly, the integration of allele specific information from eQTL and AD GWAS allowed us to unambiguously define the direction of the transcriptional changes in relation to increased AD risk for the fine-mapped genes (**Fig. 4b**, rightmost column). *PICALM* is a previously well-supported disease gene ^28^, where we found that the AD risk variant (rs10792832) was within an OCR_ABC_, and the risk allele was associated with both lower OCR signal and gene expression, which is consistent with predicted (DeepSEA) reduced DNase accessibility and H3K4me1 ChIP-seq signal in monocytes (**Fig. 4c, 4d**).

For three AD loci, moloc analyses provided support for involvement of only one among the multiple previously suggested risk associated genes at each locus: *EPHA1-AS1, USP6NL*, and *CCDC6*. The *EPHA1-AS1* locus is of particular interest as our analysis prioritized it over the *EPHA1* protein coding gene, and its 95% credible set contains a single SNP (**Fig. S17, Fig. S18**). *EPHA1-AS1* is a lncRNA gene with a previously undefined function, which we predicted to participate in immune-related pathways based on functional annotations of the co-regulated genes (**Fig. S19**).

Intriguingly, we identified three instances of colocalization between AD GWAS and microglia meta-eQTL at genetic regions not previously highlighted as AD loci, for genes *FIBP, LRRC25* and *KCNN4*, with the moloc colocalization with meta-caQTLs also observed for the latter (**Fig. 4b; Table S6, Fig. S20; Fig. S21; Fig. S22**). The known biology of all three genes is highly compatible with AD pathophysiology, with *KCNN4* having been previously considered as an AD therapeutic target ^29^. Importantly, the utilization of the generated microglia-specific regulome resources allowed us to identify AD relevant coherent regulatory units for 18 out of 21 AD loci for which were able to observe fine-mapping evidence, highlighting the importance of obtaining data from the relevant cell type.

## Transcription factor regulatory networks capture AD genetic variation

TFs are involved in the precise tuning of microglial homeostasis and are implicated in AD pathogenesis^30,31^. In parallel to pursuing the colocalization approaches and E-P annotation to link AD risk loci to genes, we also utilized chromatin accessibility to establish the TF activity in microglia and to query the identified TF targets for AD risk variant enrichment. We performed TF footprinting analysis ^23^ in microglia, as well as oligodendrocytes and GABAergic and glutamatergic neurons ^5^ for comparative analysis. PU.1 (encoded by *SPI1*), IRF6, STAT2, and NFKB2 were identified as microglia-specific TFs (**Fig. 5a**; **Table S7**), reflecting known microglial immune-related processes ^24,32–34^, with PU.1 identified as the strongest microglia-specific TF ^35^ (**Fig. 5c**). Among the 26,003 OCRs with predicted PU.1 binding, 86% (OR=16.2, p<10-16, Fisher’s exact test) matched sites identified by PU.1 ChIP-seq ^9^, further validating the *in silico* footprinting approach to detect microglial TFs.

**Fig. 5.**
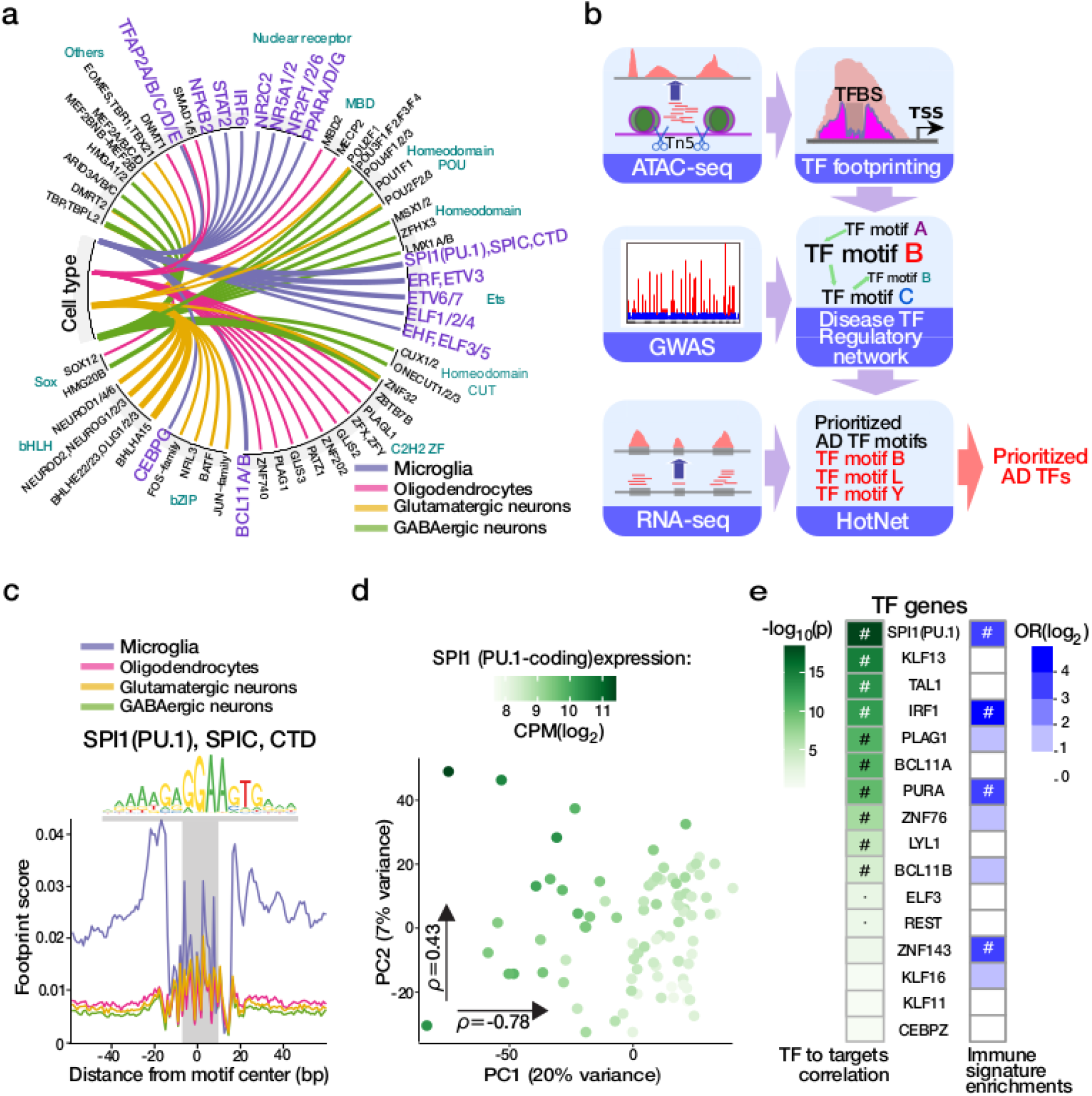
Transcription factor binding landscape in microglia integrating AD genetics. **a)** Top cell-specific TF binding events detected by TF footprinting in the microglia and three other major brain cell lineages ^5^. Line thickness indicates the fold enrichment in the highlighted cell types compared to the mean number of bound TFs in other cell types (all BH<0.05, one-sided binomial test); **b)** Schema for AD TF prioritization analysis; **c)** Aggregated footprint profile of PU.1 motif within the jointly called OCRs in the four cell populations; **d)** Principal component analysis of expression for predicted PU.1 targets genes for n=127 samples colored by expression of PU.1-encoding SPI1 gene. Spearman correlation (ρ) with each principal component; **e)** Prioritization of TFs from AD TF regulatory networks based on correlation with respective downstream target genes (shaded in green by p-value, ‘#’=BH<0.05, ‘·’=p-value<0.05). Right column: Enrichment analyses of the TF downstream target genes for immune-related gene signatures. The values represent odds ratio enrichment for immune-related signatures among all functional signatures. Significant enrichment (BH<0.05) is indicated by “#”.

Cross-regulation among groups of TFs in a given cell type defines regulatory subnetworks that underlie cellular identity and facilitate the integration of complex cellular signals. Using predicted TF binding within OCRs from footprinting analysis, we constructed a directed TF-to-TF regulatory network (TFRN) capturing the hierarchical TF regulome in microglia (**Fig. 5c, Fig. S23**). We then used AD genetic risk variants ^12^ to assign weights to the nodes in the generated TFRN ^36^ and identified subnetworks composed of 11 TF motifs jointly representing perturbed regulatory hubs in AD (**Fig. 5d**). Of the 23 TF genes corresponding to these motifs, 16 were expressed in microglia.

We prioritized TFs within this set based on the correlation of TF gene expression with the expression of their respective predicted target genes. Expression of PU.1-encoding *SPI1* had the strongest correlation with the transcriptional landscape of its 4,226 predicted target genes (**Fig. 5e, Fig. S24**). Altogether, we observed significant downstream signatures for 10 of the 16 TF genes (**Fig. 5e**, left column). Reflecting known microglial biology, the downstream target genes for three of these TFs (*SPI1*/PU.1, IRF1 and ZNF143) have a predominantly immune function as illustrated by the over-representation of immune related signatures among the enriched biological pathways (**Fig. 5e**, right column).

## Discussion

In the current study, we examined how genetic regulation of chromatin accessibility affects transcription in primary human microglia. With microglia comprising a small fraction of all brain cells, any resources generated using brain homogenate do not comprehensively capture the microglia-specific regulome. To address this gap in our knowledge, we generated multi-omics data comprising ATAC-seq, RNA-seq, and Hi-C using microglia cells isolated from 150 unique donors. We present the largest, microglia specific, meta-eQTL analysis to date and the first publicly available human microglia caQTL dataset. Incorporating Hi-C derived 3D chromosomal loop data allowed us to link accessible chromatin to target genes, leading to the identification of ∼25,000 discrete regulatory E-P units, regulating 9,890 genes. The majority of these interactions were not observed in analysis of previous data sets.

In 14 previously implicated AD risk loci, we identified disease regulatory units associated with expression of an individual gene. We confirmed previously implicated genes, *BIN1, PICALM, CD33, CASS4, ADAMTS4, INPP5D* and *APH1B*, in 7 independent loci ^37,38^. Going one step further, our approach allowed us to fine-map AD risk loci and identified 8 genes, *EPHA1-AS1, USP6NL, CCDC6, AC099524*.*1, ZNF652, MS4A4E, RABEP1* and *CLU*, as the causal genes in previously unresolved loci containing multiple candidate AD risk genes.

In the case of *EPHA1-AS1*, this locus is an example where our multi-omics approach enabled fine-mapping of a particular AD risk gene paired with the genetic regulatory mechanisms affecting its expression. Here, a genetic variant, rs11771145, comprises the 95% confidence intervals for *EPHA1-AS1* and two OCRs (peak 188003 at 143,413,799-143,414,777 and peak 188007 at 143,458,143-143,458,744, on chromosome 7) located 6kb and 51kb from the gene’s TSS, respectively. Rs11771145 is located 130bp from the peak 188003, with ATAC-seq signal at the OCR strongly (r=0.69) correlated with the expression of *EPHA1-AS1*. This SNP has the strongest association with AD at the locus in the original International Genomics of Alzheimer’s Project (I-GAP) study ^39^ and in the most recent European Alzheimer’s Disease BioBank (EADB) study ^38^. Interestingly, though rs11771145 was not included within the 95% credible interval in the AD GWAS ^12^ utilized in the colocalization analyses presented here, we still observed significant evidence linking the genetic regulation of *EPHA1-AS1* with AD etiology through the regulation of at least one regulatory element.

Importantly, while the majority of the observed colocalization were seen within previously reported AD loci, our colocalization analyses also identified three novel putative AD risk genes, namely *KCNN4, FIBP* and *LRRC25. LRRC25* regulates virally induced autophagy in myeloid cells ^38^. FIBP binds to acidic fibroblast growth factor (aFGF), which is released by astrocytes and enhances the activation of human microglia following LPS/IFN-g stimulation ^41^. Therefore, *FIBP* may be an intriguing link in the astrocyte-microglia axis of AD. *KCNN4*, on the other hand, has been extensively pursued as a therapeutic AD target due to its role in the removal of neurotoxic debris by phagocytosis ^29^. Our analysis indicates that alleles associated with decreased expression of all three of these genes are associated with increased AD risk.

By applying TF footprinting analysis we were able to identify TF regulatory networks. TFs whose regulatory neighborhood is enriched in AD risk genes were prioritized and, of those, the TF with the strongest downstream effect was PU.1 (encoded by *SPI1*). SPI1 has previously been associated with increased AD risk ^24,42^. CaSNPs were disproportionately overrepresented within PU.1 binding sites and, in 92% of these, alleles associated with lower OCR strength disrupt PU.1 binding motifs. A similar, albeit weaker, effect was observed for eSNPs. Thus, genetic variants that decrease binding of PU.1 were sufficient to interfere with the stability of chromatin accessibility. This observation, combined with the transcriptional changes associated with SPI1 expression, highlight a regulatory role for *SPI1*/PU.1 in microglia, with particular relevance to AD. We replicate previous evidence supporting PU.1 as a transcriptional factor critical to microglial contribution to AD ^24,35,42^, and further identify additional TFs implicated in AD.

Altogether, our multi-omic data set provides unprecedented insight into the regulation of microglia transcription, enabling annotation of a large number of distal regulatory elements and downstream genes. The strong enrichment of AD risk variants in microglia OCRs further establish microglia as a cell-type central to AD development. We were able to fine-map multiple AD loci, identifying not only the relevant genes but, in some cases, proposing the regulatory mechanisms contributing to disease, thus allowing further exploration of the nuances of genetic landscape contributing to particular neurodegenerative phenotypes. In conclusion, this resource provides an atlas of the human microglia regulome that can be leveraged by the scientific community to guide focused experiments in AD and other neurodegenerative diseases and to understand the impact on transcription of a particular risk locus.

## Materials and methods summary

The ATAC-seq (n=107), RNA-seq (n=127), SNP array (n=122), and Hi-C (n=5) data were generated from human brains of 150 individuals from four biobank resources (three based in New York City, NY, and ROSMAP from Rush University, Chicago, IL), including 123 autopsies and 27 biopsies. Brain tissue dissections from cortical regions were processed and subjected to FACS to isolate viable CD45+ microglia. ATAC-seq libraries were generated using an established protocol and processed through our bioinformatics pipeline ^5^, with 210,832 OCRs called via MACS2 ^43^. We applied variance component analysis to quantify the proportion of gene expression variation attributable to OCR covariance. To predict enhancer-promoter interactions, we employed the “activity-by-contact” method that is based on the combination of frequency (derived from Hi-C) and enhancer activity (derived from microglia ATAC-seq and H3K27ac ChIP-seq). We identified 24,497 E-P interactions. Transcription factors involved in the regulation of gene expression were identified by footprinting analysis in TOBIAS and modeled as a regulatory network. We then applied HotNet to find altered subnetworks containing TF motifs that are highly dysregulated between AD cases and controls based on transcriptomics and GWAS weights. We utilized the mmQTL method to identify 5,468 caQTLs in 95 samples, and 1,603 eQTLs in 101 samples. Meta-analysis utilizing summary statistics from two recently generated human microglia eQTL datasets allowed us to identify 7,302 meta-eQTLs with an effective sample size of 400. Similarly, 10,266 meta-caQTL were identified by integrating caQTLs from human macrophages (effective size =216). Functional impact of the identified caSNPs was evaluated via their effect on epigenetic states across multiple cell types in DeepSEA, as well as predicting changes to motif binding affinity in motifbreakR. To identify genes and accessible chromatin regions that share genetic regulation, colocalization analysis was performed with the coloc method, resulting in 1,457 co-regulated Gene-OCR pairs. Coloc was also utilized to detect colocalization between neurodegenerative disease predisposing genetic variants and meta-eQTLs or meta-caQTLs. Coherent units of genes and regulating OCRs relevant to AD risk were identified via multiple-trait-coloc (moloc). The detailed Materials and Methods are described in the **Supplementary Materials**.

## Supporting information

Supplementary Material and Methods, Results and Figures

## Data Availability

Data used for this analysis comes from the AMP-AD Knowledge Portal (https://adknowledgeportal.synapse.org/Explore/Studies/DetailsPage?Study=syn25671134) and is hosted on the Sage Bionetworks Synapse platform for access by qualified investigators. Data was generated from post-mortem tissue and has been de-identified according to the Synapse terms of use, and is available through the submission of a AD Knowledge Portal Data Use Certificate (https://adknowledgeportal.synapse.org/DataAccess/DataUseCertificates).

## Acknowledgments

We thank the patients and families who donated material for these studies. We thank the computational resources and staff expertise provided by the Scientific Computing group at the Icahn School of Medicine at Mount Sinai.

## Author contributions

J.F.F., V.H. and P.R. conceived of and initiated the project. J.F.F. and P.R. designed experimental strategies for omics profiling. A.W.C., B.K., D.B, C.P.K. and V.H. provided human brain tissue. J.F.F., S.R., S.P.K and Z.S. performed data generation. R.K., B.Z., J.B., G.E.H. and P.R. designed analytical strategies. R.K., J.B. and P.D. conducted initial bioinformatics, sample processing and quality control for the omics data. R.K. and G.E.H. developed the computational scheme and performed the downstream analysis. B.Z. performed the QTL analysis. J.B. performed the TF analysis. P.D. performed the Hi-C analysis. J.H., K.P.L. and T.R. provided additional QTL resources. J.F.F. and P.R. supervised overall data generation. G.E.H. and P.R. supervised overall data analysis. R.K., J.F.F., B.Z., J.B., G.E.H. and P.R. wrote the manuscript with input from all authors.

## Data and materials availability

The genotypes, omics data and metadata are available via the AD Knowledge Portal (https://adknowledgeportal.org). The AD Knowledge Portal is a platform for accessing data, analyses, and tools generated by the Accelerating Medicines Partnership (AMP-AD) Target Discovery Program and other National Institute on Aging (NIA)-supported programs to enable open-science practices and accelerate translational learning. The data, analyses and tools are shared early in the research cycle without a publication embargo on secondary use. Data is available for general research use according to the following requirements for data access and data attribution (https://adknowledgeportal.org/DataAccess/Instructions). For access to content described in this manuscript see: http://doi.org/10.7303/syn26207321. Code used throughout this study (through Fig. S16) is available upon reasonable request from the corresponding authors.

## Supplementary Materials (provided separately)

Materials and Methods

Supplementary Text

Figures S1-S24

Tables S1-S7

References (45–78)

