## Supplementary Material and Methods, Results and Figures for "Genetics of the human microglia regulome refines Alzheimer’s disease risk loci"

**Supplementary Materials Included:**

Materials and Methods

Supplementary Text

Figures S1-S24

Tables S1-S7

References (*45–78*)

### Materials and Methods

#### Sample providence and processing

All brain specimens were obtained through informed consent and/or brain donation programs at the respective organizations. All procedures and research protocols were approved by the corresponding ethical committees of our collaborator's institutions.

*Autopsies*

The autopsy brain specimens originated from brain donation programs at Rush University Medical Center/Rush Alzheimer's Disease Center (RADC) in Chicago, IL and The Mount Sinai/JJ Peters VA Medical Center NIH Brain and Tissue Repository (NBTR) in the Bronx, NY.

*NBTR*

Samples were collected at NBTR following similar parameters to MSBB–Mount Sinai NIH Neurobiobank cohort [46]. Collected autopsies were selected to represent a full spectrum of cognitive and neuropathological disease severity in the absence of discernible non-AD neuropathology. All neuropsychological, diagnostic and autopsy protocols were approved by the Mount Sinai and JJ Peters VA Medical Center Institutional Review Boards, with neuropathological assessments, cognitive, and medical and neurological status determinations performed according to previously published procedures. Postmortem brain tissue was placed in an ice-cooled insulated box and transported from the site of recovery to the Neurobiobank laboratory. A section of the frontal pole (Brodmann area 10) was dissected in the Neurobiobank laboratory, and rinsed in ice-cold sterile saline, placed in pre-cooled (4°C) MACS tissue storage solution (Miltenyi Biotec, cat# 130-100-008), and immediately refrigerated (4°C).

*RADC*

Samples were collected at RADC in Chicago, IL as part of two prospective studies of aging: the Religious Orders Study (ROS) [47] and the Memory and Aging Project (MAP) [48]. At the time of enrollment, participants had to be at least 53 (ROS) or 55 (MAP) years old and non-demented at the time of enrollment, and were required to sign an Anatomical Gift Act agreeing to donate their brain and spinal cord at the time of death. Extensive annual neuropsychologic evaluations were collected prior to death, with a structured, quantitative neuropathologic examination conducted at autopsy, described at https://www.radc.rush.edu/docs/var/variables.htm. After autopsy, the collected tissue was weighted, placed in ice-cold MACS tissue storage solution (Miltenyi Biotec, cat# 130-100-008) and shipped overnight at 4°C with priority shipping.

Biopsies

DLPFC samples in living individuals were collected as part of the Living Brain Project, approved by the Human Research Protection Program at the Icahn School of Medicine at Mount Sinai (STUDY-13-00415) from patients undergoing deep brain stimulation (DBS) procedure as part of standard-of-care treatment for Parkinson Disease treatment. Additional samples were collected from patients undergoing procedures for intracerebral hemorrhage evacuation (STUDY-18-01012A). During the regular course of either procedure, a burr hole is fashioned in the bone along the planned trajectory in order to expose the cortical surface. A cannula or sheath is then inserted into this exposed cortical surface. To pass this cannula or sheath safely, the cortical surface must first be "prepared." This entails sharply cutting the pial surface with a scalpel and cauterizing up to a cubic centimeter of tissue contained within a circular region of the cortex approximately five to ten millimeters in diameter. After making the incision in the pial surface, a microdissector is used to obtain a small, circular biopsy within the cortex that would otherwise be destroyed. In the case of patients undergoing intracerebral hemorrhage evacuation, 50% of the biopsy is sent to the pathologist for analysis and 50% is stored for this study. Following tissue dissection/isolation, all samples were placed in pre-chilled (4°C) MACS tissue storage solution (Miltenyi Biotec, cat# 130-100-008), and maintained at 4°C until dissociation and FACS. Samples kept in tissue storage solution for ≥48 hours were not processed.

**Isolation of microglia from fresh human brain specimens**

Fresh autopsy and biopsy tissue specimens were processed using the Adult Brain Dissociation Kit (Miltenyi Biotech cat.# 130107677), according to manufacturer's instructions. Following de-myelination (Miltenyi de-myelination kit, Miltenyi Biotech, cat.# 130096733) cells were incubated in antibody (CD45: BD Pharmingen, Clone HI30 and CD11b: BD Pharmingen, Clone ICRF44) at 1:500 for 1 hour in the dark at 4˚C with end-over-end rotation. RNase inhibitors (Takara Bio) were used throughout the cell prep. Prior to fluorescence activated cell sorting (FACS), DAPI (Thermoscientific) was added at 1:1000 to facilitate the separation of live from dead cells. Viable (DAPI negative) CD45 positive cells were isolated by FACS using a FACSAria flow cytometer (BD Biosciences). Following FACS, cell concentrations and viability were confirmed using a Countess automated cell counter (Life technologies).

**Genotyping and Imputation**

Genomic DNA for genotyping was extracted from frozen brain or buffy coat using the QIAamp DNA Mini Kit (Qiagen), according to manufacturer's instructions. Samples were genotyped using Infinium Psych Chip Array (Illumina) at the Mt Sinai Sequencing Core, with 571,496 SNPs retained after QC. Imputation was performed using the University of Michigan Imputation server (https://imputationserver.sph.umich.edu) utilizing the HRC reference panel. Only 7,723,699 SNPs with MAF >0.05 were retained for caQTL and eQTL analyses.

### Chromatin Accessibility analyses

*Generation of ATAC-seq libraries and sequencing*

ATAC-seq reactions were performed on 120 samples (113 autopsies and 7 biopsies) using an established protocol [49] with minor modifications. In brief, 75,000 sorted cells were centrifuged at 300 g for 10 min at 4°C. Pellets were resuspended in lysis buffer (10 mM Tris-Cl pH7.4, 10 mM NaCl, 3 mM MgCl, 0.1% IGEPAL CA630) by pipetting 20 times. Following resuspension, samples were centrifuged at 500 g for 10 min at 4°C. Pellets were resuspended in transposase reaction mix (25 µL 2x TD Buffer (Illumina, cat.# FC-121-1030) 2.5 µL Tn5 Transposase (Illumina, cat.# FC-121-1030) and 22.5 µL Nuclease Free H2O) on ice. Reactions were incubated at 37°C for 30 min and then purified using the MinElute Reaction Cleanup kit (Qiagen, cat.# 28204), eluting in 10 µL of buffer EB. Following purification, library fragments were amplified using the Nextera index kit (Illumina, cat.# FC-121-1011) as described previously [49]. Library quality was determined by analysis on Tapestation D5000 ScreenTapes (Agilent technologies, cat.# 5067-5588). Following amplification, libraries were resolved on 2% agarose gels and fragments ranging in size from 100-1000bp were excised and purified (Qiagen Minelute Gel Extraction Kit, Qiagen cat.# 28604). Before sequencing, libraries were quantified with the Qubit dsDNA HS assay kit (Invitrogen, cat.# Q32851) and using quantitative PCR (KAPA Biosystems, cat.# KK4873). Fragment sizes were estimated using Tapestation D5000 ScreenTapes (Agilent technologies, cat.# 5067-5588) and libraries were sequenced on Hi-Seq2500 (Illumina) obtaining 2x50 paired-end reads or the NovaSeq6000 platform (Illumina) obtaining 2x100 paired-end reads.

*ATAC-seq quality control and further processing*

*Alignment of reads.* Sequenced reads were delivered by the sequencing facility already demuxed and with adaptors trimmed. FASTQ files were matched to the respective samples based on pooling IDs and barcodes. The one to one match and file integrity was confirmed using MD5 checksums.

Reads were subsequently aligned to the hg38 reference genome with the pseudoautosomal region masked on chromosome Y with the STAR aligner (v2.7.0e) and the following parameters:

```
--alignIntronMax 1
--outFilterMismatchNmax 100
--alignEndsType EndToEnd
--outFilterScoreMinOverLread 0.3
--outFilterMatchNminOverLread 0.3
```

For STAR and other java programs, java v1.7.0 was used. The alignment yielded a BAM file for each sample consisting of mapped paired-end reads sorted by genomic coordinates. From these files, reads that mapped to multiple loci were removed using samtools, duplicated reads were removed with PICARD (v2.2.4; http://broadinstitute.github.io/picard) and, finally, reads mapping to the mitochondrial genome were also removed. Quality was assessed using Qualimap, fastqc, and phantompeakqualtools. For the latter and other programs in python, v2.7.14 was used.

*Genotype calling.* Genotypes were called using GATK (v3.5.0). In brief, the following steps were performed: (1) indel-realignment; (2) base score recalibration; and (3) joint genotype calling across all samples for variants with a phred-scaled confidence threshold ≥ 10. All clustered variants, variants in ENCODE blacklisted regions of the genome, and variants not in dbSNP v151 were not considered. Read depth was not used as a filtering criterion. Finally, only variants with minor allele frequencies (MAF) ≥ 25% were retained. For genotype file processing, bcftools, vcftools and plink were used. The genotype concordance amongst samples was quantified using both the fraction of concordant genotype calls and the kinship coefficient from KING v1.9. Both approaches gave comparable outcomes, and both indicated an unambiguous separation of samples from the four different individuals.

*Peak Calling and Read Quantification for Quality Control.* Peak calling was performed using MACS v2.1 [43] as previously described [5]. In short, we merged samples with the same disease phenotype (AD, Controls, and Other, which includes non-AD neurodegenerative, neuropsychiatric, and a number of other phenotypes associated with poor health states) into one BAM-file. The three resultant bam files were then subsampled to a uniform depth and used as input for peak calling.

*Gender determination of samples.* Three different metrics were used to assess the gender of the samples: 1) The rate of heterozygotic genotyping calls on the X chromosome outside the pseudoautosomal regions. For this, variants with MAF < 5% were discarded. In samples from male individuals, a high heterozygosity rate potentially indicates sample contamination or an incorrect gender. 2) The read counts of OCRs adjacent to FIRRE and XIST, which predominantly show chromatin accessibility in samples from female individuals. 3) Read counts in OCRs identified on the Y chromosome outside the pseudoautosomal region. No gender mismatches were identified using these three metrics.

*Quality control and further processing.* For each sample, the following quality control metrics were used: the total number of initial reads; the number of uniquely mapped reads; the fraction of reads that were uniquely mapped; further metrics from the STAR aligner; the duplication and insert metrics from Picard; the rate of reads mapping to the mitochondrial genome; the PCR bottleneck coefficient (PBC), which is an approximate measure of library complexity estimated as uniquely mapped non-redundant reads divided by the number uniquely mapped reads; the normalized strand cross-correlation coefficient (NSC) and the relative strand cross-correlation coefficient (RSC), which are metrics that use cross-correlation of stranded read density profiles to assess sample quality independently of peak calling; and, finally, the fraction of reads in peaks (FRiP), which is the fraction of reads that fall in detected peaks, the fraction of reads in only blacklisted peaks, and the ratio between these two metrics (for these metrics the consensus set of peaks was used) (**Table S1**).

We excluded libraries that had a low FRiP (<20%), had a low final read count (<5 million reads), visually were outliers in clustering, or looked to have outright failed when inspecting the bigWig track. After QC steps, only one sample was left from a Parkinson disease focused biobank, and it was subsequently left out, thereby leaving 107 samples for downstream analyses (**Table S1**).

The samples with the same neurodegenerative disease related phenotype were subsequently subsampled and merged, creating 3 BAM-files (AD, Controls, and Other neurodegenerative and neuropsychiatric disease phenotypes) with a uniform depth of 240 million paired-end reads. Using these BAM-files, bigWig files were created using bedtools, bedGraphToBigWig, wigToBigWig, and wiggleTools. Peaks were called with the same parameters as for QC. A consensus set of peaks was subsequently created requiring a peak to be called in one or more of the merged BAM-files. After removing peaks overlapping blacklisted genomic regions, 210,832 peaks remained. Next, read counts of the individual 107 non-merged samples within these peaks were quantified, again, using the same parameters as for quality control.

*Comparison with other brain based ATAC-seq datasets.*

Peaks generated via MACS2 for each external dataset (**Table S2)** with CPM>1 in at least 20% samples per dataset, were merged into a combined peak set (404,101 unique OCRS), and the peaks were re-quantified jointly for all external datasets similarly to how our microglia dataset was processed. Keeping only OCRs outside blacklisted regions, and present at CPM>1 in at least 5% of samples in two or more datasets, 359,377 OCRs were retained. Dataset identity, gender, and QC metrics ("fracReadsInNonBlacklistedPeaks" and "picard_meanGcContent") were included in voom modelling after TMM normalization, with the resulting expression matrix residualized for the gender and QC metrics. Pairwise Spearman correlation between samples in the residualized expression matrix was visualized using MDS.

**Transcriptome analysis**

*Generation of RNA-seq libraries and sequencing*

Where available, 100,000 cells were sorted into 1.5 ml low-binding Eppendorf tubes containing Extraction buffer, a component of the PicoPure RNA Extraction kit (Arcturus, cat# KIT0204). and incubated at 42°C for 30 min while shaking at 850 rpm. Samples were stored at -80°C prior to RNA extraction according to manufacturer's instructions. This included an RNase-free DNase treatment step (Qiagen, cat.# 79254). Samples were eluted in RNase-free water and stored at -80°C until preparation of RNA-sequencing libraries using the SMARTer Stranded Total RNA-seq Pico Kit v1(Takara Clontech Laboratories, cat.# 635005) or v2, according to manufacturer's instructions. RNA-seq libraries were quantified by quantitative PCR (KAPA Biosystems, cat.# KK4873) and library fragment sizes estimated using High Sensitivity Tapestation D1000 ScreenTapes (Agilent, cat.# 5067-5584). RNA-seq libraries were subsequently sequenced on NovaSeq 6000 (Illumina) machines yielding 2x50, 2x100, or 2x150 bp paired-end reads.

*RNA-seq processing*

The generated RNA-seq libraries were processed by RAPiD (ASHG ref), a locally implemented standard pipeline. TRIMMOMATIC [50] was utilized to remove adapters and discard low quality reads. Pair end reads were aligned to the human reference genome GRCh38/hg38 via STAR [51], implementing

WASP module [52] to correct for allelic bias in mapping due to individual patients' genetic variants for 125 samples with available SNP genotyping data. The generated BAM files contained splice junction, and transcript/isoform level quantification was performed with RSEM [53], summarized for gene level analyses. Quality control metrics were generated via Picard (v2.2.4; http://broadinstitute.github.io/picard). The identities of the samples were confirmed by comparing the genetic variants called from RNA-seq reads between the tested samples, comparing with genetic variants from ATAC-seq data, as well as imputed genotypes from samples with available SNP array data. Sex identity was validated via expression of *XIST* (female-specific) versus *RPS4Y1* (male-specific) gene.

The total of 165 samples with available RNA-seq data included isolated microglia populations from 146 patients. For the further analyses we retained one sample per patient, keeping only samples isolated from the brain cortex, containing either the entire $CD11b^{+}CD45^{+}$ microglial compartment (134 samples), or the larger microglial compartment (10 samples). One patient had two cortical samples from two brain regions, of which the one with the largest number of reads (consistent with the larger numbers of input cells) was retained. To perform quality control for the remaining 143 samples, we considered a number of metrics, including low amount of starting material and low ratio of uniquely mapped reads. Based on these, we removed 11 samples as these had less than 50% of uniquely mapped reads (more than two times of standard deviations from the distribution of that measure). Most of these removed samples (n=7) were obtained from biopsies, which, due to the very limited amount of tissue, had lower yield of sorted cells available for RNA-seq libraries compared to those available from autopsies.

For the remaining 132 samples, we calculated the difference of mean correlation of the given sample with the rest of the dataset. We identified five samples whose mean correlation of all pairs of samples within the dataset was more than three times higher than the standard deviation calculated upon all pair's correlation, and removed them from further analyses. The remaining 127 samples were utilized for further downstream analyses. All RNA-seq QC metrics are summarized in **Table S1**.

*Comparison with other relevant RNA-seq datasets.*

Neuronal, astrocytes, and oligodendrocytes data, processed utilizing the standard in-house RAPiD pipeline, were downloaded from ARCHS4 resource (https://maayanlab.cloud/archs4/data.html) [54]. Data for monocytes [55], microglia (Raj) [8], and microglia (Gaffney) [7] samples were processed similarly, with transcript level counts obtained from Kalisto [56] collapsed at gene-level. Only genes with CPM>1 in >10% of the samples were retained. Gene counts were normalized using TMM method, and only samples with >5M library size were kept. MDS analysis was applied to each dataset, and any sample with SD>2.5 or 3 (according to visual examination of distribution) for the top 2 dimensions were removed. After the QC steps the following cohorts of samples were collected: Astrocytes n=289, Neurons n=300, Oligodendrocytes n=12; Monocytes n=230, Microglia (Gaffney) n=117, Microglia (Raj) n=255, Microglia (Roussos) Autopsy n=103, Microglia (Roussos) Biopsy n=24.

### Transcription factor analysis

To estimate chromatin occupancy by transcription factors, we performed footprinting analysis using TOBIAS (v.0.10.1) [23] as detailed in the following.

*Motif selection*

We downloaded human TF binding motifs from the CIS-BP 1.02 meta-database [57], which contained 3,059 motifs. Because many transcription factors are associated with more than one motif, and since the transcription factors belonging to the same transcription factor family generally share binding motifs, we had to determine which motif is the most representative one for each transcription factor. To do so, we applied majority vote approach: The candidate motifs for that given transcription factor were extracted and all pairwise similarities between these candidate motifs were calculated by TomTom [58] based on the following command line parameters [59]: `tomtom -dist kullback -query-pseudo 0.1 -target-pseudo 0.1 -text -min overlap 0 -thresh 1`. Outputted similarity scores were log-transformed and summed for each candidate motif representing the transcription factor, and the motif showing the lowest score was chosen. This resulted in 431 motifs, representing 798 transcription factors.

*Estimation of chromatin occupancy by transcription factors*

The footprinting analysis was performed separately for (1) comparison of microglia samples with other brain regions and (2) microglia samples only. In the former case, we started the analysis with the merged set of all OCRs detected in microglia as well as glutamatergic neurons, GABAergic neurons, and oligodendrocytes [5], i.e. 386,146 OCRs (6.42% of the genome). Each cell type was represented by one BAM file, each consisting of 354 million reads that were randomly selected from the corresponding samples. In the latter case, given the focus on Alzheimer's disease, we split microglia samples to AD patients, AD controls, and "other" (representing non-AD neurodegenerative diagnosis) groups, thus increasing the sensitivity to capture AD-specific binding events. Each of these groups was represented by one BAM file, consisting of 240 million reads (again randomly selected from the corresponding samples) and profiled in 207,248 microglial OCRs (3.54% of the genome). For both analyses, we ran the TOBIAS module ATACorrect to correct for Tn5 insertion bias in input BAM files, followed by TOBIAS ScoreBigwig to calculate footprinting scores across OCRs. Then, TOBIAS BINDetect combined footprinting scores with the information of transcription factor binding motifs to evaluate the individual binding positions of each transcription factor and determine whether a given position was bound by a given transcription factor or not for each condition, i.e. cell type and brain region. Finally, TOBIAS PlotAggregate was used to visually compare the aggregated footprints for select motifs.

*Calculation of TFRN networks and prioritization of TF motifs*

We assembled a TF regulatory network (TFRN) in microglia (using microglia samples only) capturing TF-to-TF interactions among the 431 TF motifs (nodes). Directed connections (edges) were defined by the presence of actively bound transcription factor binding sites in the proximal regulatory regions (< 3 kb from TSS) of 798 TF genes. Then, we used this network as an input of the HotNet algorithm 36 to find altered subnetworks containing TF motifs that are highly dysregulated based on GWAS and are topologically close on an interaction network. Gene-level AD GWAS weights were calculated by MAGMA 1.07b [60] using default settings but removing the broad MHC region due to its extensive linkage disequilibrium and complex haplotypes [61], as well as the ApoE region due to its extreme signal and complex haplotypes [12]. In the case of multiple TF genes per one TF motif, we set the weight to be the highest -log10(*P*-value) and applied the Sidak method for multiple testing corrections [62]. Only TF motifs that participated in at least 50 TF-to-TF interactions were kept.

*Prioritization of TF genes represented by the same TF motif*

Because each TF motif can be represented by multiple TF genes, we needed to recognize phenotypically causal ones. Therefore, for each TF motif, we detected all genes that have a given motif actively bound in the promoter region (within 3 kb from TSS). We subsetted the expression matrix for those genes and calculated its principal components. Lastly, we quantified the Pearson correlation coefficient of the first principal component and the expression of all TF genes representing a given TF motif. We kept only significantly correlated TF genes using Bonferroni adjusted *P*-value threshold of 0.05.

*Integrating caQTLs with DeepSea epigenome predictions*

DeepSEA trained a deep neural network to predict the presence/absence of an epigenetic annotation given only the local genome sequence [21,22] (https://humanbase.readthedocs.io/en/latest/deepsea.html). The effect of a genetic variant was estimated by evaluating the probability of an epigenetic annotation given the reference sequence and comparing it to the probability given the alternative allele. For each variant DeepSea returns the log of the probability difference. For each of 8,786 caQTL SNPs located within the regulated OCR and which had posterior inclusion probability > 1%, the DeepSea scores were obtained for each of 2,002 assays from ENCODE and the Epigenomics Roadmap Project in the DeepSea resource. The direction of the DeepSea score for each SNP was re-coded based on direction of the caQTL regression coefficient so that a positive value indicates that DeepSea and the caQTL coefficient have the same predicted direction. For each assay, the mean of the re-coded DeepSea effect size was evaluated. A positive mean indicates concordance between the DeepSea predictions for the assay and the caQTL analysis from the current work. The set of 2,002 mean values was then converted to z-scores by subtracting the mean and dividing by the standard deviation. A p-value was then computed for each z-score. The p-values computed across assays roughly follow the theoretical null distribution except for a set of assays from myeloid lineage samples.

**Generation of Hi-C libraries and sequencing**

Hi-C data was generated from five fresh postmortem human brain tissue using the in situ Hi-C protocol [63] with the following modifications. FACS sorted cells were fixed with 0.5% formaldehyde for 10 min and then quenched with 0.125 M glycine for 5 min at RT. Cross-linked tissue was then placed on ice for a further 15 min to quench crosslinking completely. Samples were centrifuged at 800 g for 10 min at

4°C and pellets resuspended in lysis buffer (0.32 M Sucrose, 5 mM CaCl2, 3 mM Mg(CH3COO)2, 0.1 mM EDTA, 10 mM Tris-HCl, pH 8, 1 mM DTT, 0.1 % Triton X-100, 1x Roche cOmplete mini EDTA-free protease inhibitor tablet (Roche cat.# 4693159001) to isolate cross-linked nuclei.

Approximately 1 M crosslinked nuclei were thawed on ice, washed with ice-cold 1x CutSmart buffer (New England Biolabs (NEB), cat. B7204S) and split into 4 aliquots to generate technical replicate libraries per sample, with 250,000 nuclei per library. Nuclei were pelleted at 2,500 g for 5 min at 4°C, and resuspended in 342 µl 1x CutSmart buffer and then conditioned with 0.1% SDS at 65°C for 10 min. Nuclei were immediately placed on ice and the SDS quenched with 1% Triton X-100. Chromatin was digested with 100 U of the 4 base pair cutter MboI (NEB, cat.# R0147L) overnight at 37°C with shaking at 400 rpm. MboI was heat-inactivated at 65°C for 20 min, and then nuclei were cooled down on ice. MboI cut sites were end-labeled with biotin by adding 52 µl of biotin fill-in reaction mix (15 µl of 1 mM biotin-14-dATP (Jena Bioscience, cat.# NU-835-BIO14-L), 1.5 µl each of 10 mM dCTP, dGTP, dTTP (Sigma-Aldrich, cat.# DNTP10-1KT), 10 µl of 5 U/µl Klenow DNA Pol I (NEB, cat.# M0210L), 22.5 µl of 1x CutSmart buffer) and shaking at 37°C for 1.5 h at 400 rpm. Blunt ended sites were proximity ligated by adding 948 µl ligation reaction mix (150 µl of 10x T4 DNA ligase buffer (NEB, cat.# B0202S), 125 µl of 10% Triton X-100, 15 µl of 10 mg/ml BSA, 10 µl of 400 U/µl T4 DNA ligase (NEB, cat.# M0202L), 648 µl ddH2O) and rotating tubes, end-over-end, at RT for 4 h. Nuclei were reverse crosslinked with 100 µl proteinase K (10 mg/ml) overnight at 65°C.

Proximity ligated DNA was purified through phenol:chloroform extraction and sodium acetate/ethanol precipitation. Purified DNA was sheared using a Covaris S220 sonicator to generate a peak size of 400 bp with the following settings (peak incident power: 140 W, duty cycle: 10%, cycles per burst: 200, time: 55 sec). Biotin-labeled ligation junctions were purified with Dynabeads MyOne Streptavidin C1 beads (ThermoFisher, cat.# 65001) by incubating for 1 hr at RT. Illumina compatible libraries were prepared from the sonicated and streptavidin bead immobilized DNA using the NEBNext Ultra II Library prep kit (NEB, cat.# E7645L), following manufacturer's instructions, by amplifying libraries for 6-10 PCR cycles. Libraries were purified by 2-sided size selection (300-800 bp) using Ampure XP beads (Beckman Coulter, cat.# A63881). All libraries were analyzed on a TapeStation using Agilent D5000 ScreenTapes (Agilent technologies, cat.# 5067-5588), and quantified using the KAPA Library Quantification Kit (KAPA Biosystems, cat.# KK4873) prior to sequencing. Uniquely barcoded Hi-C libraries were pooled and deep sequenced on the Illumina NovaSeq S4 platform (Illumina) obtaining 100 bp paired-end reads.

**Hi-C data analysis**

Hi-C data were aligned using the HiC-Pro strategy [64]. Briefly, paired-end reads were mapped independently to the human genome hg38 using bowtie2 in stringent mode with parameters ("--very-sensitive -L 20 --score-min L,-0.6,-0.2 --end-to-end") [65]. Then, the chimeric reads that failed to align were trimmed after ligation sites (MboI "GATCGATC") and mapped to the genome. All the aligned reads from both ends were then merged based on read names and mapped to MboI restriction fragments using hiclib package [66]. Next, self-circles, dangling ends, PCR duplicates, and genome assembly errors were discarded. Samples of the same cell type were merged. We binned the interaction matrix at different resolutions and corrected it with iterative correction (ICE) for downstream analysis [66]. All Hi-C QC metrics are summarized in **Table S1**.

### Variance component modeling of gene expression

A variance component analysis was used to examine how much gene expression variability could be correlated to patterns of chromatin covariance. To perform this analysis, we followed an implementation suggested by a previous report [67]. First, we modeled negative binomial distribution of RNA-seq count data by a variance stabilizing transformation (vst; varistran R package) [68]. Then, for each gene represented by vst-normalized vector g, we considered the following variance component model:

$$Y_g' = N(0, P,\ OCR_{EP}\sigma_{ocr}^2 + U\sigma_u^2)$$

where $OCR_{EP}$ is the sample-sample covariance matrix of chromatin accessibility in promoter (OCRs overlapping region within 1kb from the transcription start site) and enhancer regions (OCRs overlapping region within 1-100kb from the transcription start site), and $U\sigma_u^2$ is the noise term. The values of $\sigma_{ocr}^2$ and $\sigma_u^2$, were estimated by the average information restricted likelihood estimation (AIREML; gaston R package) [69]. We used residualized count matrices of OCRs where the effect of technical covariates was regressed out. Importantly, this approach does not model the relationship of each gene to its own promoter/enhancer OCRs but to the overall status of all enhancers/promoter OCRs across all chromosomes. Permuted analyses (100 runs) were done by shuffling the sample labels for the vst-transformed expression matrix for the 88 samples with both ATAC-seq and RNA-seq data available.

### Prediction of enhancer-gene interactions

We used Activity-by-contact model (ABC, v.0.2) [13] to construct a comprehensive regulatory map of enhancer-promoter (E-P) interactions in neuronal and non-neuronal cell types of the two investigated brain regions (STG and EC). This model requires: (1) contact frequency between putative enhancers and promoters of regulated genes; and (2) enhancer activity data. Contact frequency matrices were generated from microglia Hi-C generated from five post-mortem human brains (current study). Here, different neocortical regions (dorsolateral prefrontal cortex, orbital frontal cortex, and anterior prefrontal cortex) were profiled across multiple donors aged 34-103 years. Enhancer activity data were represented by the cell type and brain region specific ATAC-seq signal (current study) and previously published H3K27ac ChIP-seq data [4]. In accordance with the authors' directions, we filtered out predictions for genes on chromosome Y and lowly expressed genes (genes that did not meet inclusion criteria in our RNA-seq dataset). We used the default threshold of ABC score (a minimum score of 0.02) and the default screening window (5MB around the TSS of each gene).

### QTL analyses

*eQTL and caQTL detection*

To identify caQTL and eQTL in the fresh microglia, we utilized mmQTL, a flexible statistical package to control for population structure [15]. Genetic relatedness matrix, used as an input, was constructed using GCTA Software Tool (`--autosomes --make-grm` ) with genome-wide normalized genotypes after removing 15 regions with high LD. For each of genes or annotated OCR, variants in cis-region, defined as 500kb and 50kb around explored feature were tested for the association with the normalized gene expression or ATAC-seq signal (described below). Multiple test correction was applied on both locus-level and genome-level through the BH method. FDR was controlled at 5%.

The expression and ATAC-seq data as utilized for QTL analyses was processed as below. We utilised Bayesian Information analyses to identify technical variables which need to be corrected for, and in combination with critical demographic and clinical measures (age, gender, neuro-degenertaive disease status) comprised pre-selected variables utilised in latent variable testing. The count matrices were normalized via trimmed tean of M values (TMM) method [70], and transformed via voom modeled with the pre-selected variables. The number of PEER factors were determined by generating a range of latent factors (from 1 to 30) utilizing peer R package [71] while considering the pre-selected variables of interest. Each individual set of identified latent factors, in conjunction with the pre-selected variables, was used to create a residualized matrix, which was then utilized by fastQTL [72] to identify QTLs correcting for genetic ancestry with 10 MDS components generated in the genotyped dataset (after removing 15 genomic regions with high LD). The numbers of QTLs (FDR < 0.05) were calculated in each analysis, and the number of latent variables producing the largest number of detected QTL before leveling off was utilized for the mmQTL analyses.

*Meta-QTL analysis*

To improve the statistical power in QTL detection, we previously developed a novel meta-analysis method, mmQTL, and applied it on both eQTL and caQTL detection. mmQTL provides a flexible meta-analysis pipeline to integrate QTL among datasets, either individual-level or summary results. Briefly, it firstly performed QTL detection in each of the datasets, and assembled the QTL signal, of which non-significant variants were used to estimate covariance due to phenotypic correlation. A mixed linear model was then used to estimate parameters, and RE2 [73] was applied to test statistical significance. In real application on eQTL, summary results of eQTL from primary microglia samples collected from Lopes *et al*. and Young *et al*., and together with the eQTL signal from the dataset created in this study were provided to perform meta-analysis. As to caQTL, we downloaded caQTL summary results from Alasso *et al*., [16] which contained four macrophage datasets, one is naive, and three in stimulation status. Based on the accompanied ATAC-peak annotation, we extracted 64,740 OCR peaks uniquely overlapped with our ATAC-seq peaks (at least 1 base pair overlapping), and mmQTL was applied to leverage caQTL signal among five datasets on these overlapped ATAC-seq peaks. The identified 7,380 meta-caQTLs were combined with the 2,889 microglia-only caQTLs for the 140,285 OCRs present in the microglia dataset only.

*Estimation of cell type contribution to meta-caQTL signal*

We applied M-value, which is a Bayesian method to calculate the posterior probability measuring QTL existence in each study of a meta-analysis [18], to show the caQTL sharing and cell-type specificity. For each ATAC-seq peak with significant caQTL, we extracted the top-variant which has the most significant caQTL evidence, and estimated the M-value in five datasets. Considering the small sample size in both macrophage and microglia datasets, and multiple datasets available for macrophage data (generated in cells with four different activation states), we used a looser cutoff 0.8 to determine support for a caQTL. For macrophage caQTLs we required an M-value of at least 0.8, observed in at least two of the macrophage datasets, to have considered the caQTL present in macrophages.

*Estimation of effective sample size for meta eQTLs and caQTLs*

Considering that in the meta QTL detection, we included the dataset with samples from diverse populations, an effective sample size was estimated based on the reported variance of QTL signals. eQTL signals were detected only in n=93 European individuals in Young *et al*. 7, which is then used as baseline to compare with the reported variance in meta-eQTL results, and effective sample size was calculated as $n*median(\sigma_{Young}{}^{2}/\sigma_{meta}{}^{2})$. The same pipeline was used to estimate effective sample size for meta-caQTL analysis, with the only difference that each of four macrophage caQTL results was used as reference, and the final estimator was calculated as the average of four effective sample sizes. For caQTL meta-analyses the effective size was 216 samples, and for eQTL meta-analyses the effective sample size was 400.

### Partitioned heritability analysis

We partitioned heritability for OCRs, caSNPs, and eSNPs to examine the enrichment of common variants in neuropsychiatric traits with stratified LD score regression (v.1.0.0) [74] from a selection of GWAS studies (**Table S3**). Briefly, for the OCRs a binary annotation was created by marking all HapMap3 SNPs [75] that fell within the peak, while for eSNPs and caSNPs, PP scores were used for the continuous annotation. LD scores were calculated for the overlapped SNPs using an LD window of 1cM using 1000 Genomes European Phase LD reference panel [76]. The enrichment was determined against the baseline model. To enable comparisons of the regression coefficients across traits with a wide range of heritabilities, we chose to normalize by the per-SNP heritability and named this adjusted metric the “normalized heritability coefficient”.

To estimate the contribution of colocalized eQTL-caQTL to traits, we first calculated a colocalization posterior probability, which measures the probability that the eQTL-caQTL share the same causal variants, based on MaxCPP annotation in eQTL and caQTL. The constructed joint posterior probability was then used as continuous annotation to run partition heritability analysis.

### COLOC and MOLOC colocalization

To evaluate the relationship between molecular QTL, we used COLOC, an R package to conduct colocalization analysis. The output Z statistics from meta-analysis was used as input for COLOC [26,27].

The phenotypic variance was set to 1, as we have normalized the summary results before meta-analysis. We applied an extension of COLOC, MOLOC [26,27], evaluating the colocalization of multi-omics data, summary results from meta eQTL, meta caQTL and GWAS summary results were used as input. The parameter was set to be default in implementation of COLOC and MOLOC. For COLOC analyses we considered colocalizations with pp4 > 0.5 as significant [7], and pp15 > 0.5 for MOLOC [27].

**Joint Fine-mapping**

The joint fine-mapping analysis was performed following Hormazdiar [77], where posterior inclusion probabilities (PIP) from the same genetic variant are multiplied together for two traits to compute the colocalization posterior probability (CLPP). A CLPP > 0.01 was considered significant consistent with a widely used cutoff motivated by empirical analysis and simulations [15].

**Evaluation of disruptive effect of caQTL on TF binding**

We applied motifBreakR to annotate the disruptive effect of caQTL on TF binding [78]. Briefly, fine-mapped caQTL variants located in ATAC-seq peak and in the 95% credible set were mapped to 630 known TF motifs (JASPAR2016). MotifBreakR calculated the relative entropy for the reference allele and alternative allele within annotated OCRs with the setting threshold = 1e-4, method = “ic”. We only kept the strongly affected TFBSs, calculated the allele’s binding ability difference, and compared it with the reported beta value in caQTL results. To determine what TFs are more likely to be affected by caQTL, we first sampled the same number of ATAC-seq peaks, from which we randomly chose the same number of variants as that in real fine-mapped caQTL variants, and the number of variants affecting TF binding ability were compared with that in real data. Fisher’s Exact test was used to calculate significance using the mean number of 100 random samples among genetic variants within the peaks, and FDR was controlled at 5% by BH method.

### Supplementary Results

*AD snps in ABC enhancers*

The biological and clinical relevance of the microglia regulatory landscape can be interrogated by examining the distribution of the identified OCR-Gene links in relation to fine-mapped neuro-degenerative disease risk variants. We focused on Alzheimer's disease due to consistent and significant enrichment of variants within microglia OCRs, and, in particular, those with high ABC scores. Remarkably, 6,428 distal $OCR_{ABC}$ (>20kb from the nearest TSS) contain 20 SNPs within 95% credible intervals (PP>0.01) from eight different AD loci identified by Jansen et.al while the 97,513 of the equally distant OCRs with low ABC scores contained none of the SNPs within these same 95% credible intervals (OR=318, $p=8.9 \times 10^{-25}$, Fisher's exact text) (**Fig. 4a, Table_S6**). The eight loci with SNPs included within $OCR_{ABC}$ the genes linked via ABC included previously suggested as potential etiologic candidates: *ADAMTS4, INPP5D, USP6NL, PICALM, CASS4*, among others.

*AD snps in promoters*

Genetic variants from the 95% credible intervals from seven AD loci (three of which overlapped with the eight ABC colocalized loci, above) were located in OCRs within 3kb of a gene's TSS. Of the seven OCRs, five were unambiguously proximal to genes encoding *INPP5D, APH1B, SCIMP, CD33* and *CASS4*. A promoter OCR at chromosome 17 (at 58.3Mb) was located near the TSS for both *TSPOAP1* and *TSPOAP1-AS1* genes. At another locus on chromosome 17 (5.3Mb) a fine-mapped AD snp (rs75511804, PP=0.084) was located within a promoter of the SCIMP gene, while another fine-mapped SNP (rs61481506, PP=0.057) was within an OCRABC linked to *RABEP1*.

*Moloc colocalization analyses*

A particularly useful approach for fine mapping of disease loci is the application of colocalization methods, such as coloc, as a means of integrating AD GWAS with eQTLs to identify genetic variants contributing to both disease etiology and regulation of a particular gene. Since we also have caQTL information, we applied an extension of the original coloc method, multiple-trait-coloc (moloc) (*32*), to combine summary statistics from AD GWAS (*4*) with the generated microglia meta-eQTL and meta-caQTL datasets. Using this approach, we obtained statistical support for colocalization within six reported AD loci (**Fig. 4b, Table S6**), providing coherent units of transcriptional regulation relevant to the etiology of AD. For three AD loci on chromosomes 2 (127Mb) and 11 (86Mb), the link included *BIN1* and *PICALM* genes for which there is ample prior support for a role in AD. At the locus on chromosome 14 (at 92Mb), both *SLC24A4* and *RIN3* genes had comparable moloc support for involvement in AD via the same implicated OCR. However, at loci on chromosomes 7 (143Mb) and 10 (11Mb and 59Mb), moloc analyses provide support for involvement of only one among the multiple previously suggested risk associated genes at each locus: *EPHA1-AS1, USP6NL*, and *CCDC6*, respectively (**Fig. 4b, Table_S6**). Importantly, the integration of allele specific information from eQTL and AD GWAS allowed us to unambiguously define the direction of the transcriptional changes in relation to increased AD risk for the fine-mapped genes (**Fig. 4b,** rightmost column).

Of particular interest, of course, is the ability to resolve previously ambiguous etiologic gene prioritization for a number of AD GWAS loci. One example where our data is able to provide an important insight is a locus on chromosome 16 near *PLCG2*. While *PLCG2* has been implicated in AD risk via rare nonsynonymous genetic variants, a recent study by Bellengues and colleagues highlights the presence of two independent AD loci in the region. One locus is driven by common variants upstream of the *PLCG2* gene (rs12446759 is the index SNP), while the second is driven by rare variants within the P*LCG2* gene (rs72824905 is the index SNP). We observe significant colocalization between Jansen *et al* AD GWAS risk variants and AC099524.1 eQTL (pp4=0.52), where rs12444183 is the index eSNP (PP=0.88) and also has the most significant evidence for association with AD in the region in Jansen *et al* GWAS results (p=1.41*$10^{-5}$). AC099524.1, with a proposed name *BCALM*, is a potentially intriguing AD candidate gene, described as a non-coding lncRNA gene involved in Ca2+ signaling downstream from B-cell Receptor (BCR) in B-cells, the function shared with *PLCG2*, though *AC099524.1* failed to influence *PLCG2* expression in experimental models.

Among the loci where the combination of various modalities was particularly informative in elucidating the contribution of genetic variants to AD, was the region at chromosome 17 (143.4Mb) where most prior publications highlight EPH (ephrin) Receptor A1 (*EPHA1*) as the putative risk gene. In this region we observe evidence from moloc for three-part colocalization between AD and *EPHA1-AS1*, and two OCR regions (**Table S6, Fig. S17, Fig. S18)**. A SNP in the region, rs11771145, is an eSNP for *EPHA1-AS1* and a caSNP for nearby Peak_188003 (PP = 0.99) for both (**Fig. S17**). *EPHA1-AS1* expression and Peak_188003 strength are highly correlated (ρ=0.69) (**Fig. S18**). Interestingly, this eQTL/caQTL pair was the only one among 1,457 eQTL/caQTL colocalized pairs that demonstrated FDR significance (q<0.10) for Causality Inference Test for SNP>OCR>Gene effect mediated by rs11771145 (**Fig. S18a)**. The major allele G (MAF=0.61) is associated with AD, and is associated with both lower *EPHA1-AS1* expression and Peak_188003 strength. Rs11771145 is the index SNP in the largest AD GWAS/GWAX study to date, has the highest PP in IGAP 2013 study (0.36), has PP of 0.062 in Marioni et al. (AD or family history of AD phenotype), but is not within the 4 SNPs comprising 95% credible set in Jansen et. al. study. The function of *EPHA1-AS1*, a non-coding lincRNA, is not well established, and its expression is not correlated with the expression of *EPHA1* (ρ=0.07). The genes whose expression is positively correlated with *EPHA1-AS1* (Spearman rank test p-value <0.01) are, among other biological processes, enriched for genesets involved in inflammatory and anti-viral responses (**Fig. S19**). Altogether the available genetic evidence supports *EPHA1-AS1* as the AD etiologic gene in the region, whose dysregulation is mediated by reduced chromatin accessibility at Peak_188003. Moloc analyses also provide significant statistical link between AD and *USP6NL* at chromosome 10 (11.6Mb) (*ECHDC3* and *AL512631.1* had also been candidates at this locus), and with CCDC6 on the same chromosome at 60.0Mb locus (where *ANK3* gene had also been considered a plausible AD risk gene).

Apart from identifying disease relevant genes, our data can be utilized to better understand how genetic variants contribute to AD for well supported disease genes. In the case of *PICALM1* (phosphatidylinositol binding clathrin assembly protein), the identity of the *PICALM1* as an AD associated risk gene is well established (*36*). However, the addition, via moloc analyses, of caQTL information highlighted the importance of a particular OCR (Peak_30728). Two genetic variants, rs10792832 and rs3851179, in high LD with each other (r2 = 0.97), comprise the 95% critical interval for both *PICALM* eQTL and Peak_30728 caQTL, and have comparable evidence for association with AD. AD risk allele G at rs10792832 was associated with both lower OCR signal and lower gene expression, and was predicted by DeepSEA analyses to be significantly associated (FDR < 0.05) with reduced DNase accessibility and H3K4me1 ChIP-seq signal.

*Novel Putative AD genes*

The majority of colocalization observations supported the contribution of genes located within previously identified AD GWAS loci, suggesting that these approaches capture the disease relevant relationships within the framework of genome-wide significance supported genomic regions. Intriguingly, we identified three instances where colocalization methods provided significant support for a contribution to AD for genes not previously located within established AD loci.

In the first case we observed eQTL-AD GWAS colocalization is Fibroblast Growth Factor (Acidic) Intracellular Binding Protein (*FIBP*) at chromosome 16 (81.7Mb). FIBP is widely expressed, and its loss was associated with overgrowth syndrome and potential tumor predisposition. It is an intriguing AD candidate as it selectively binds to acidic fibroblast growth factor aFGF, which was reported as enhancing the pro-inflammatory phenotype in microglia following LPS or IFN-γ stimulation via FGFR2 IIIb. The locus includes 21 genetic variants associated with AD with p-value $<1*10^{-5}$, each of which are also within the 95% credible interval for FIBP eQTL (**Fig. S20**).

Another gene linked to AD via the eQTL-AD GWAS colocalization encodes for leucine rich repeat containing protein 25 (*LRRC25*) on chromosome 19 at 18.4Mb (PP4=0.93), presenting another putative novel locus for AD. The gene's function is very compatible with AD pathophysiology, being a key negative regulator of type I interferon signaling by targeting RIG-1 for autophagy in myeloid cells, and located within a C3aR-centered network observed in AD. Three SNPs within the locus have reached the significance threshold for suggestive disease loci (p-value $<1*10^{-5}$), with rs3859570, located 3kb upstream of the *LRRC25* TSS, reaching nominal significance of $8.4*10^{-7}$ (**Fig. S21**). Two of the three most significant AD SNPs are eSNPs for LRRC25, with PP of 0.26 and 0.32.

In the third instance, we observed evidence for AD_eQTL_caQTL colocalization via MOLOC at chromosome 19 (23.8Mb) for genetic variants shared between Jansen AD GWAS with *KCNN4* and Peak_82668, as well as *RP11-15A1.3* with Peak_82668 and Peak_82669. We also observe eQTL-caQTL colocalization between these two genes and each of the two peaks. Two of the SNPs within the region have evidence for association to AD with p-value $<1*10^{-5.}$ Two other SNPs, which are within

95% credible interval for KCNN4 eQTL, have AD association p-value $<1*10^{-4}$ (**Fig. S22**). *KCNN4/KCa3.1* ($Ca^{2+}$/calmodulin-activated K+ channel), has been linked to downstream activation of p38 MAPK following LPS stimulation and contributing to microglia activation and nitric oxide-dependent neurodegeneration, whereas *RP11-15A1.3* is a lincRNA gene implicated in bone mineral density and osteosarcoma, and all of the 75 SNPs within 95% credible interval for RP11-15A1.3 eQTL have AD association p-value > 0.05.

Altogether, out of 29 AD loci from Jansen *et al*, we can provide evidence supporting a predominant contribution for a single gene for 14 of the loci, while the evidence was equivocal in supporting contribution of multiple genes for an additional two loci (**Fig4b).** Importantly, our approach provides supporting evidence for three novel putative AD risk genes, further extending our knowledge of the genetic basis of AD. In addition, for many of the identified genes we can provide putative regulatory mechanisms for how disease associated risk alleles contribute to the genes' dysregulation.

### Supplementary Discussion

Application of footprinting analyses allowed us to detect instances of TFs bound within individual microglial OCRs and led to the generation of directed TF regulatory networks. TFs whose regulatory neighborhood is enriched in AD risk genes were prioritized as critical for AD, and, of those, the TF with the strongest downstream effect was PU.1 (encoded by *SPI1*), with a known, critically important, role in microglia. The genes downstream of SPI1 activity are involved in multiple immune processes, and higher expression of PU.1 was associated with increased AD risk, while knock-down of PU.1 in the microglial cell line, BV2, alleviated the pro-inflammatory response. A particular relevance of SPI1 in the regulome of microglia was observed independently when we pursued the functional impact of caSNPs. These were most disproportionately overrepresented within PU.1 binding sites with the allelic changes significantly affecting the TF's binding ability, and, in about 90% of these, the alleles associated with lower OCR strength also disrupted the binding motifs. A similar, albeit weaker effect was observed for eSNPs. Thus, genetic variants that decrease PU.1 binding within the OCRs were sufficient to interfere with the stability of chromatin accessibility for a large number of the affected regions. This observation, combined with the transcriptional changes associated with SPI1 expression, highlight a particularly critical role *SPI1*/PU.1 has on the regulatory landscape in microglia, with particular relevance to the etiology of AD.

A critical aspect of regulome analysis is annotation of accessible chromatin regions with respect to regulated genes. By applying the recently described Activity-By-Contact (ABC) framework, incorporating 3-D chromosomal structure detected by Hi-C, to link OCRs to genes' promoters, we identified 24,497 E-P links, providing regulatory annotation for 52% of the expressed microglial genes and 6% of the identified OCRs. The functional relevance of these ABC detected E-P links was supported by: a) stronger relationship between the implicated OCRs and the linked genes' expression and the proximal promoters' strength; b) higher probability of a SNP within $OCR_{ABC}$ to be an eSNP; c) higher probability of an eSNP to be located within an OCR with high ABC; and d) higher probability of a caSNP regulating the OCR with a high ABC score to also regulate the gene's transcription. However, the most dramatic evidence for the biological relevance of the OCRs with E-P links was observed by the relationship between the E-P links and AD. First, we observed a much greater enrichment of AD risk genetic variants within $OCR_{ABC}$, with normalized heritability coefficients from LD score regression analyses increasing from 2.5, for all detected microglia OCRs, to 18 for those with high ABC score. Second, the localization of fine-mapped AD risk SNPs was limited to distal (Distance to a TSS20kb) OCRs with high ABC scores. Thus, the E-P links do indeed identify open chromatin regions with strong contribution to transcription of downstream genes in a manner consistent with the microglial contribution to disease. Importantly, most of the human microglia regulome landscape is not captured by data generated in a broader cell compartment. For example, the majority of the E-P links identified via the ABC method in microglia were not detected in a broad glia population, with over 80% of the microglia-detected pairs in the current study being novel.

The utilization of colocalization methods (coloc and moloc) have shown to be effective means to fine-map GWAS loci by highlighting genes whose regulatory landscape is significantly consistent with the pattern of disease genetic association within a previously defined locus. Multiple-trait colocalization analysis of both caQTL and eQTL with the AD GWAS summary level results allowed for unambiguous fine-mapping of *EPHA1-AS1, USP6NL* and *CCDC6* in AD loci with previously uncertain causal gene identification. Importantly, in each of these cases we also identified one or more regions of co-regulated accessible chromatin, allowing for definition of the regulatory mechanisms contributing to the disease relevant modulations of the genes' expression. At two other loci, BIN1 and PICALM, we confirm the support for the previously established risk genes while identifying the relevant regulatory elements, whereas at the *RIN3/SLC24A4* locus we observed comparable evidence for both genes. We also observed colocalization for AD GWAS and eQTLs for *PLCG2, ABI3* and *CD33* loci, and between AD GWAS and caQTLs for *MS4* and *CASS4* loci. Thus, the total number of previously implicated AD loci with colocalization support via transcription or regulatory chromatin accessibility observations in microglia is ten. Six of these were previously supported by the colocalization analyses with microglia eQTL in the latest large AD fine-mapping study, which also presented microglia eQTL colocalization findings for *TSPNL14* and *PTK2B* not observed by us. The direction of the changes in the genes transcription associated with the increased AD risk as established by our analyses is largely consistent with the estimations of Jansen *et al.*: higher levels of *BIN1*, and lower levels of *PICALM, SLC24A4* and *CASS4* are associated with AD risk, with the only disagreement observed for *CD33* potentially explained by splicing perturbations. The high replication of the prior microglia QTL-AD colocalization results by Schwartzentruber *et al.* in our analyses provides confidence in the validity of the colocalizations observed by us. Therefore, it is particularly intriguing to consider the observed colocalization between AD GWAS and microglia eQTL for the three genomic regions outside the established AD loci (PP4 >0.5 for colic and/or PP15>0.5 for moloc). In all three cases the eSNPs have demonstrated association with AD with p-values between $5x10^{-8}$ and $1x10^{-5}$, thus not reaching genome-wide significance threshold according to the genetic evidence alone. The three putative AD risk genes include *FIBP, LRRC25* and *KCNN4*, all of which are excellent functional candidates for involvement in

AD etiology. Amongst these genes, the first binds to acidic fibroblast growth factor (aFGF), associated with a pro-inflammatory microglia phenotype, and the second regulates virally induced autophagy in myeloid cells; therefore both genes being very plausible candidates for AD etiology. Alleles associated with lower levels of both of these genes are associated with the increased AD risk, consistent with reported effect of *LRRC25* deficiency enhancing antiviral response in a monocyte cell line. For the *FIBP* the reduced expression with AD risk is of interest since the aFGF, a binding target of *FIBP,* is released by astrocytes enhances the activation of human microglia following LPS/IFN-γ stimulation, thus making *FIBP* an intriguing link in the astrocyte-microglia axis of AD etiology and pathophysiology.

However, the latter of the three new putative genes, *KCNN4*, may be a particularly intriguing putative AD risk gene. It has been extensively pursued as a therapeutic AD target due to its potential for selective inhibition of neurodegenerative aspects of microglia functions while preserving the neuroprotective phagocytic ability to remove neurotoxic debris. There are two *KCNN4* eSNPs, rs8104447 and rs8111664. For the former, allele C is positively associated with the gene's expression (β=0.544, p=0.00070, PP=0.74), and is negatively associated with AD risk (β=-0.0084, p=9.6x10-5), with a similar behaviour for allele A at the second SNP: positive association with the gene's expression (β=0.535, p=0.00058, PP=0.25), and is negative association with AD risk (β=-0.0084, p=9.6x10-5). For this gene the evidence for colocalization come from moloc analyses, thus also providing the identity of the accessible chromatin region genetically coregulated with the gene's transcription and disease risk: Peak_82668 at chr19 between 43,779,867 and 43,780,115) whose middle point is located within the first intron of the gene, 1,266bp from the TSS). Allele C at rs8104447 is positively associated with that OCR (p=1.5x10-6, PP=0.0210), and allele A at rs8111664 is positively associated with same peak strength (p=1.5x10-6, PP=0.0210). Thus, the AD risk alleles which are associated with the lower levels of KCNN4, which is contrary to the therapeutic approaches primarily considered for *KCNN4* focusing on the inhibition of the gene's activity for the AD treatment.

707 **Supplementary Figures:**

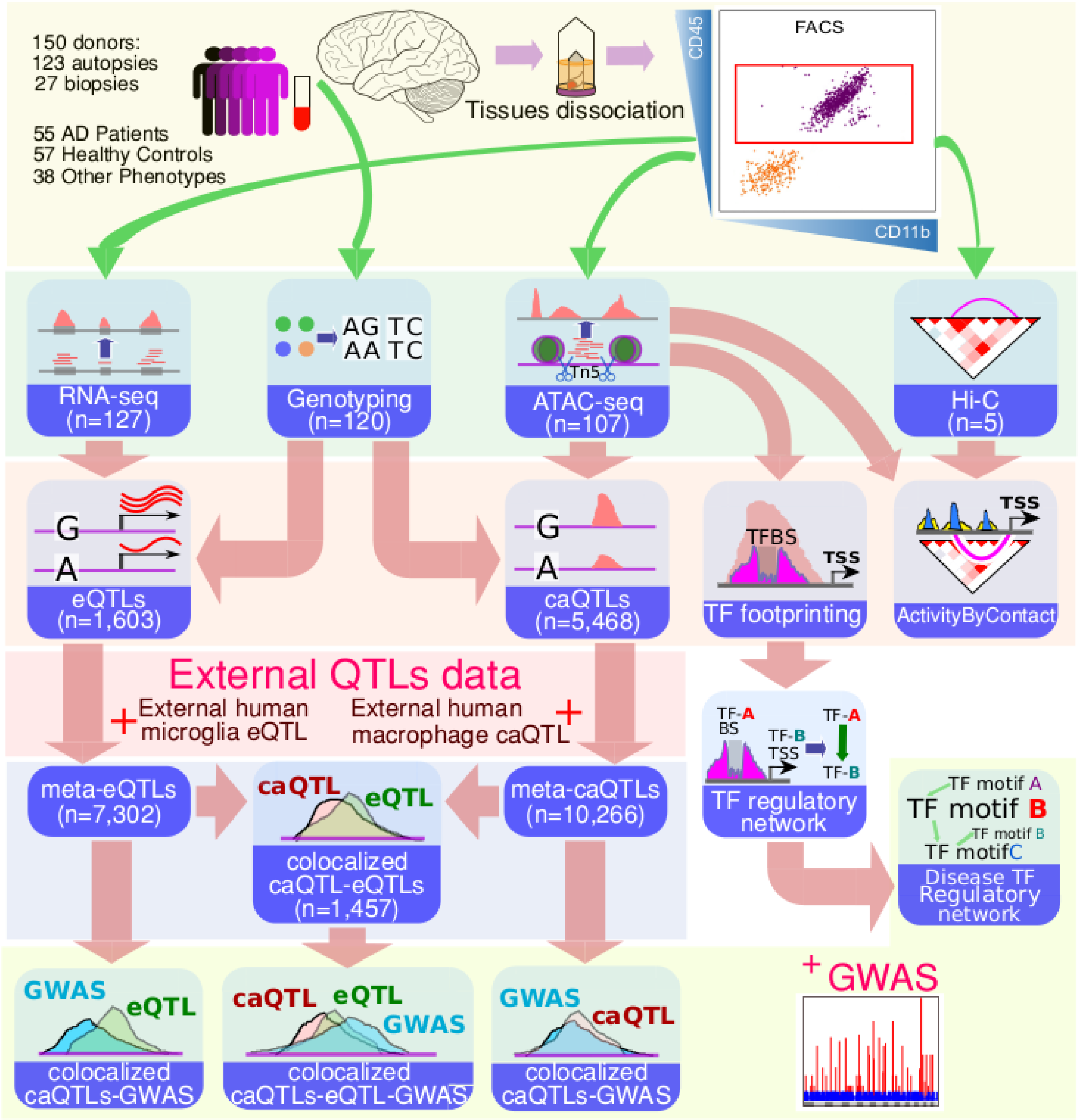


708 **Fig. S1.**

709 **Schematic outline of data generation and integrative analyses**.

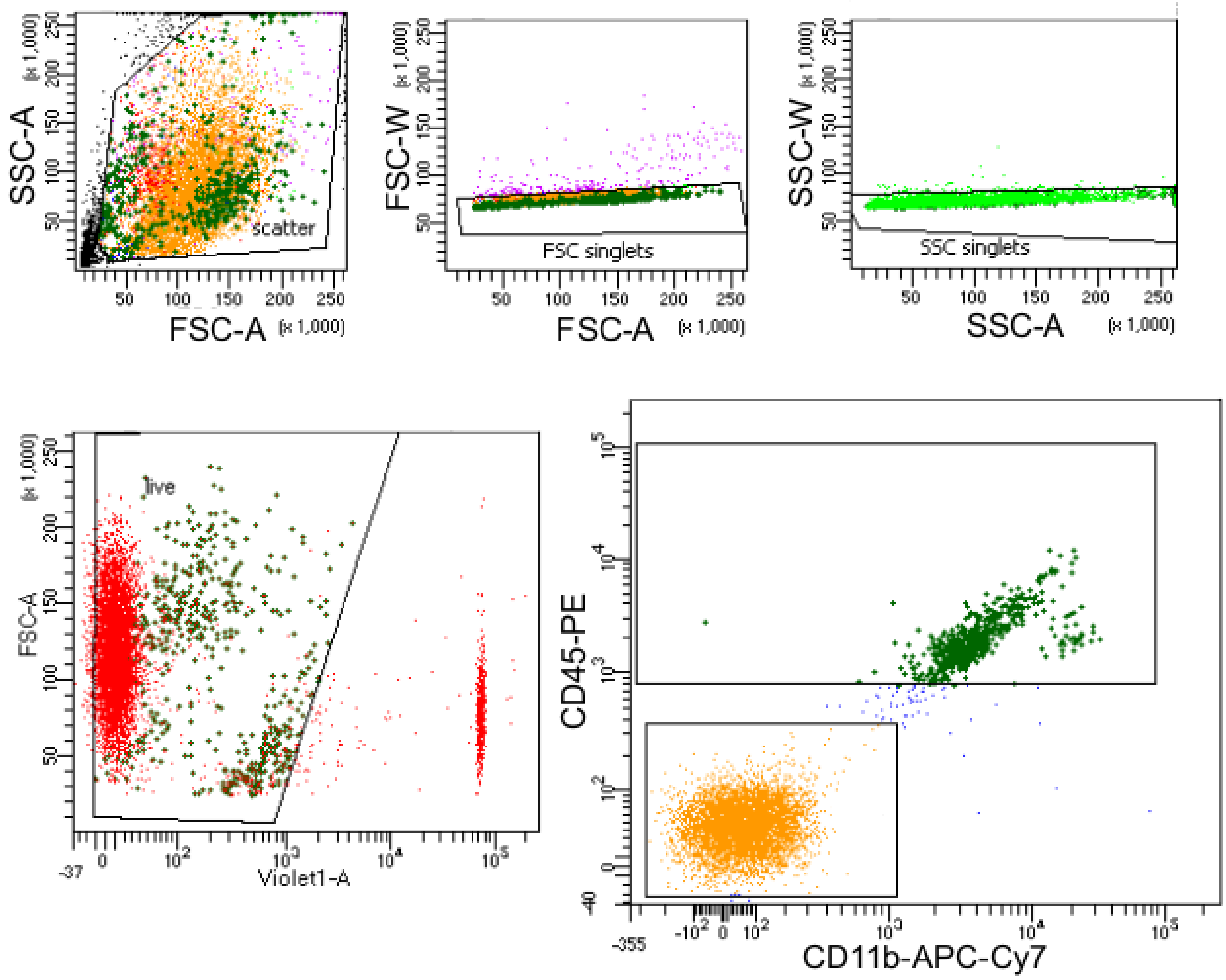


Tube: ATACseq

| Population | #Events | %Parent | %Total |
|---|---|---|---|
| All Events | 10,000 | #### | 100.0 |
| scatter | 8,705 | 87.1 | 87.1 |
| FSC singlets | 8,442 | 97.0 | 84.4 |
| SSC singlets | 8,368 | 99.1 | 83.7 |
| live | 7,320 | 87.5 | 73.2 |
| cd45- | 6,748 | 92.2 | 67.5 |
| cd45+ | 508 | 6.9 | 5.1 |

**Fig. S2.**

**FACS gating of fresh microglia**. The gating strategy for a representative sample targeting single live (DAPI-) CD45$^{+}$ microglia.

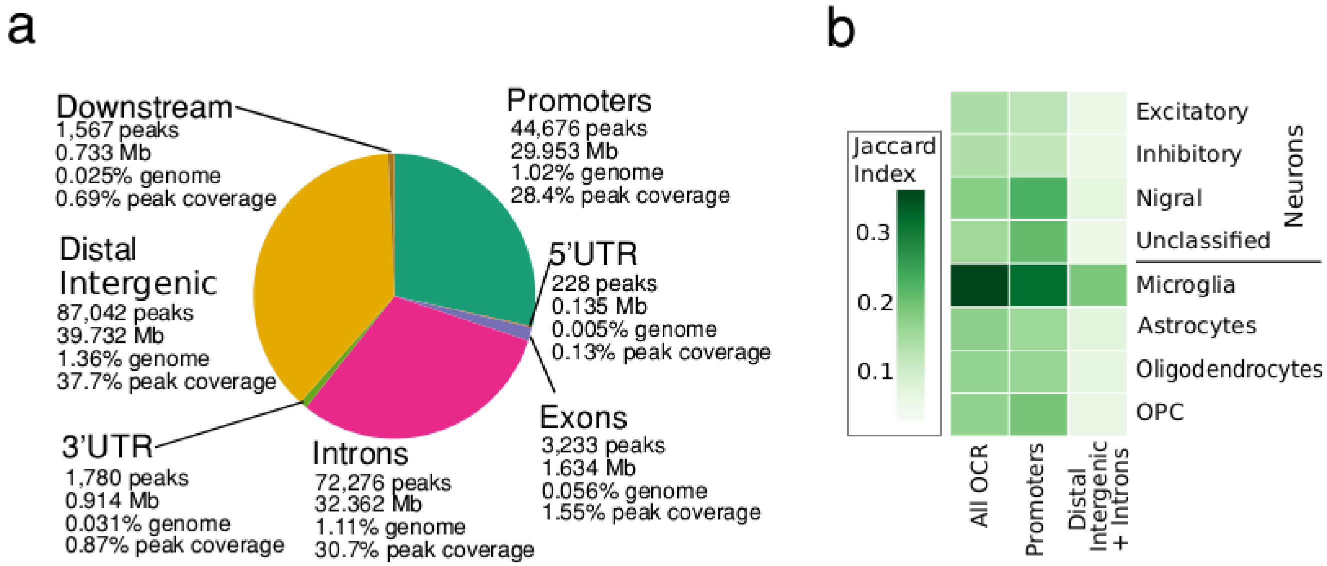


**Fig. S3.**

**Annotation of microglia OCR**. **a)** Annotation of the 210,832 OCRs detected in microglia ATAC-seq data computed using ChIPseeker R package. **b)** Comparison of human microglia OCRs to cell specific OCRs from single cells [6].

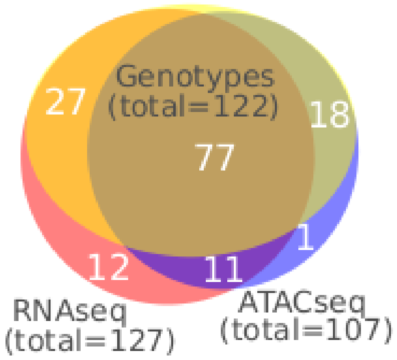


**Fig. S4.**

**Distribution of samples with ATC-seq, RNA-seq, and genotyping data**. Overlap of the high quality ATAC-seq, RNA-seq, and genotyping data for 146 donors with available ATAC-seq or RNA-seq data.

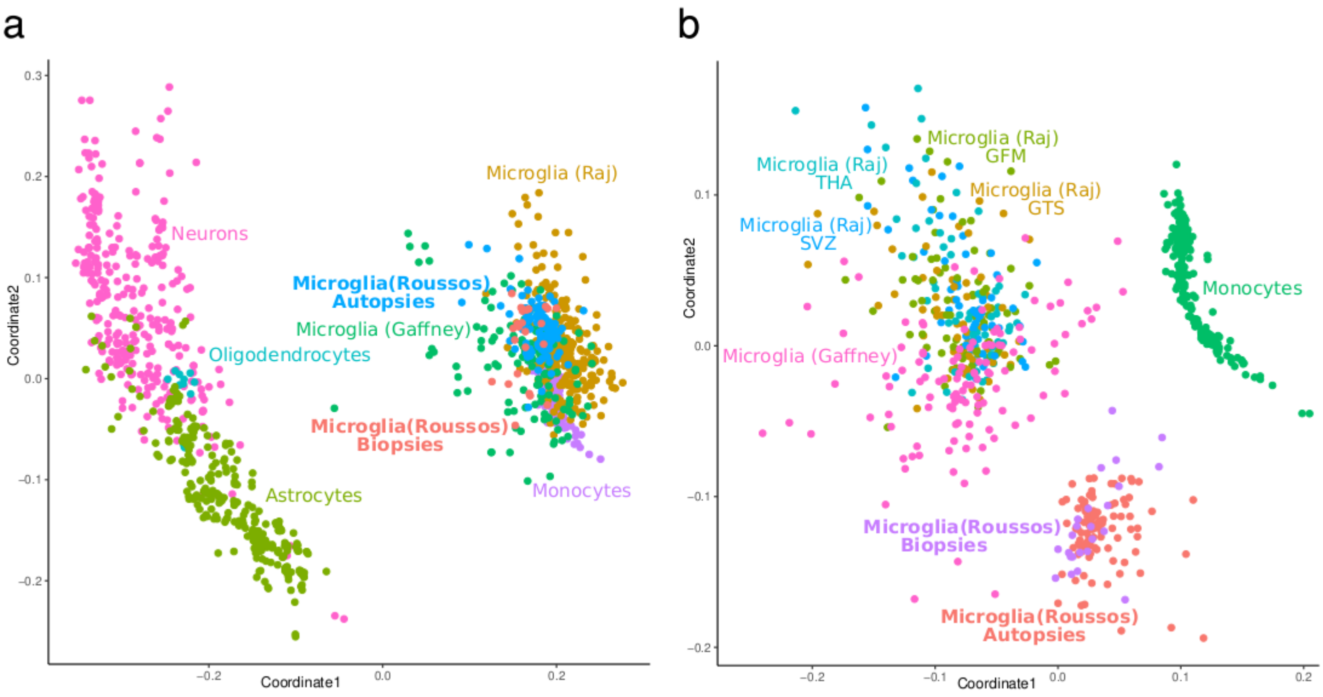


**Fig. S5.**

**Comparison of human microglia RNA-seq dataset to other available microglia datasets.** We applied multidimensional scaling (MDS) to expression data processed by the standard RAPiD pipeline for the two microglia transcriptomics datasets whose eQTLs were utilized in meta-eQTL analyses, as well as other relevant cell types. **a)** Comparisons include other brain-derived populations and b**)** only the three microglia and one monocyte datasets.

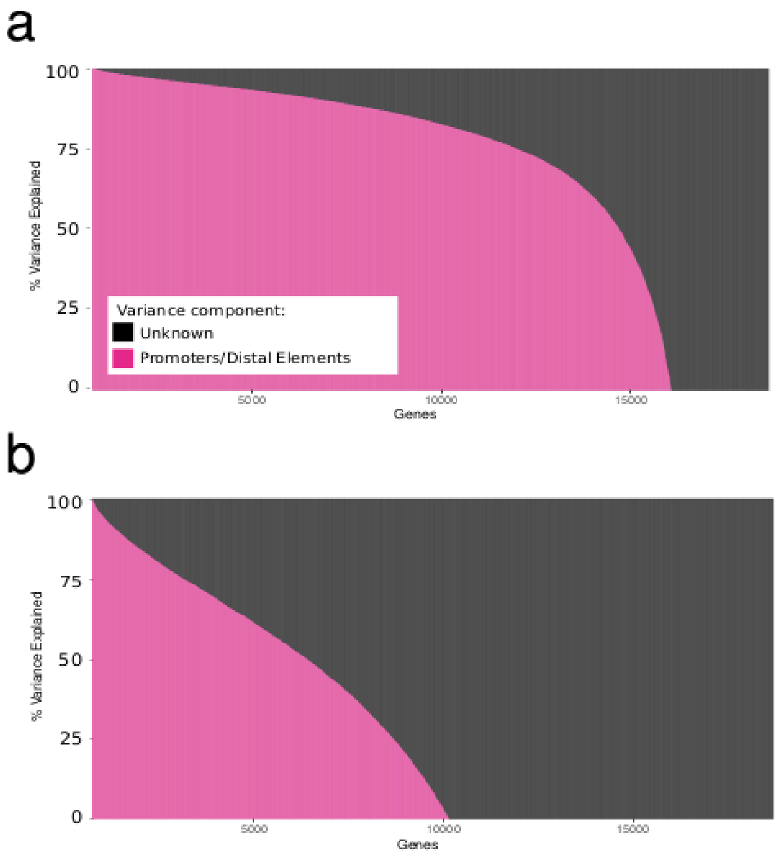


**Fig. S6.**

**Covariation of gene expression and chromatin accessibility landscapes.** The fraction of gene expression variance explained by promoter or distal enhancer OCRs across 18,856 tested genes is indicated for **a**) observed and **b**) permuted results. Genes are sorted based on the fraction of variants explained by promoter or distal elements. Permuted analysis was performed by shuffling sample labels in the RNA-seq expression matrix.

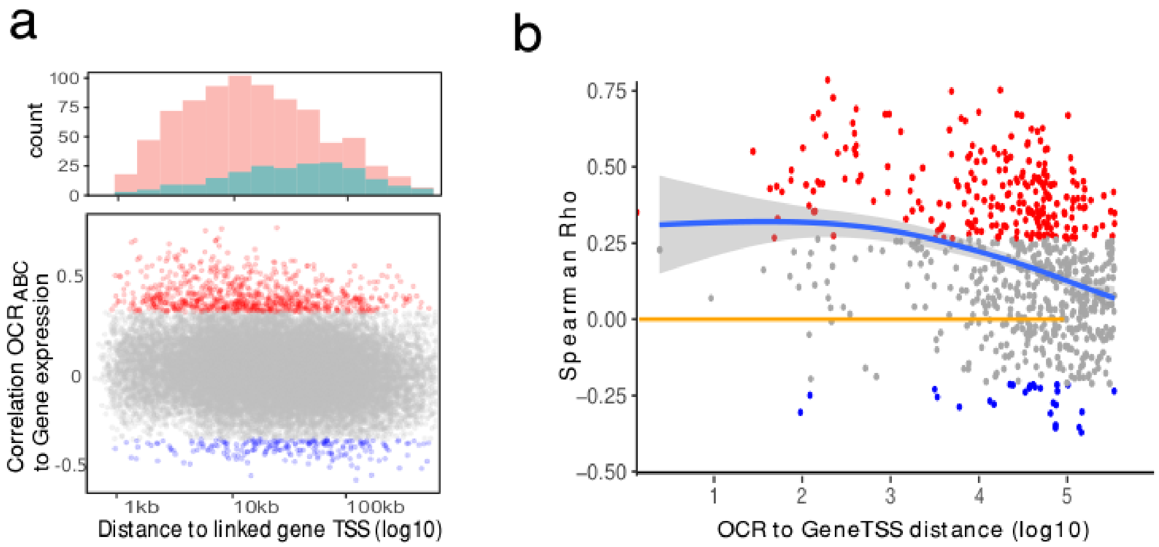


**Fig. S7.**

**Gene-OCR correlations for genes with detected E-P interactions. a)** Spearman correlation between OCR and linked gene shown for 24,497 $OCR_{ABC}$-Gene pairs plotted against the distance between the OCR and gene. More pairs show positive correlation (red, 815 pairs at FDR<5%) than negative correlation (blue, 218 pairs at FDR<5%). OCRs with positive correlation tend to be closer to the gene's TSS than OCRs with negative correlation (top panel). **b)** The same correlation versus distance plot for 1,457 colocalized caQTL-eQTL pairs represent both enhancing and repressing regulatory relationships.

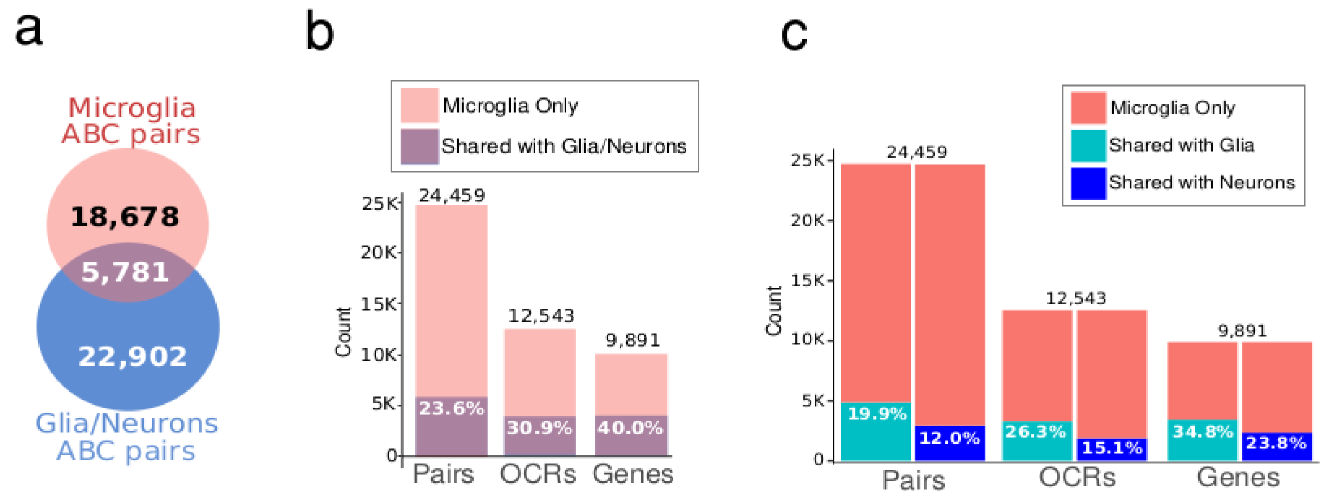


**Fig. S8.**

**Comparison of ABC derived E-P links from microglia, and in broad neuronal and glial brain fractions.** Majority of $OCR_{ABC}$-Gene pairs identified in human microglia are unique to microglia and have not been detected among $OCR_{ABC}$-Gene pairs generated in broad glial and neuronal datasets. **a)** Overlap between the 24,459 $OCR_{ABC}$-Gene pairs identified in microglia, and neuronal or broad glial cells derived 28,683 $OCR_{ABC}$-Gene pairs which include OCRs shared with microglia (out of the total of 70,778 $OCR_{ABC}$-Gene pairs detected in neurons or broad glia); **b)** The bar plots represent the distribution of the unique (i.e. detected only in microglia and not in neurons or broad glia) versus shared (present in both microglia and either broad glia or neurons) E-P interactions; **c)** Sharing of ABC E-P links identified in microglia versus neuronal (NeuN+) or non-neuronal (NeuN-) brain cell populations.

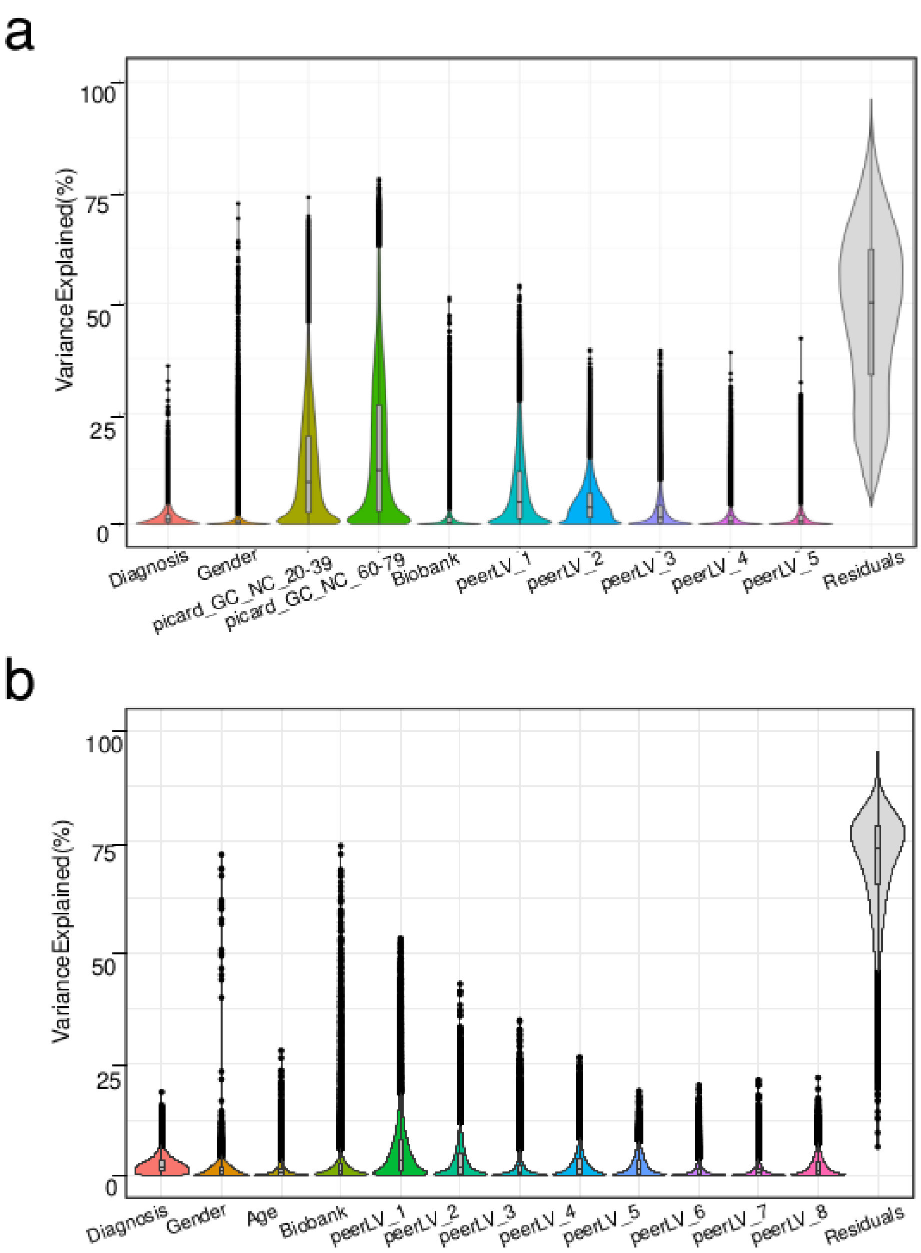


**Fig. S9.**

**Exploration of variables utilized in generating residualized matrices of chromatin accessibility and transcriptional data for QTL analyses. a)** Variance Partition analyses of variables corrected for in ATAC-seq derived OCR data utilized for caQTL detection. Selected variables included important demographic or clinical features, as well as covariates identified as important via BIC analyses; **b)** Variance Partition analyses of variables corrected for in ATACseq derived OCR data utilized for eQTL detection;

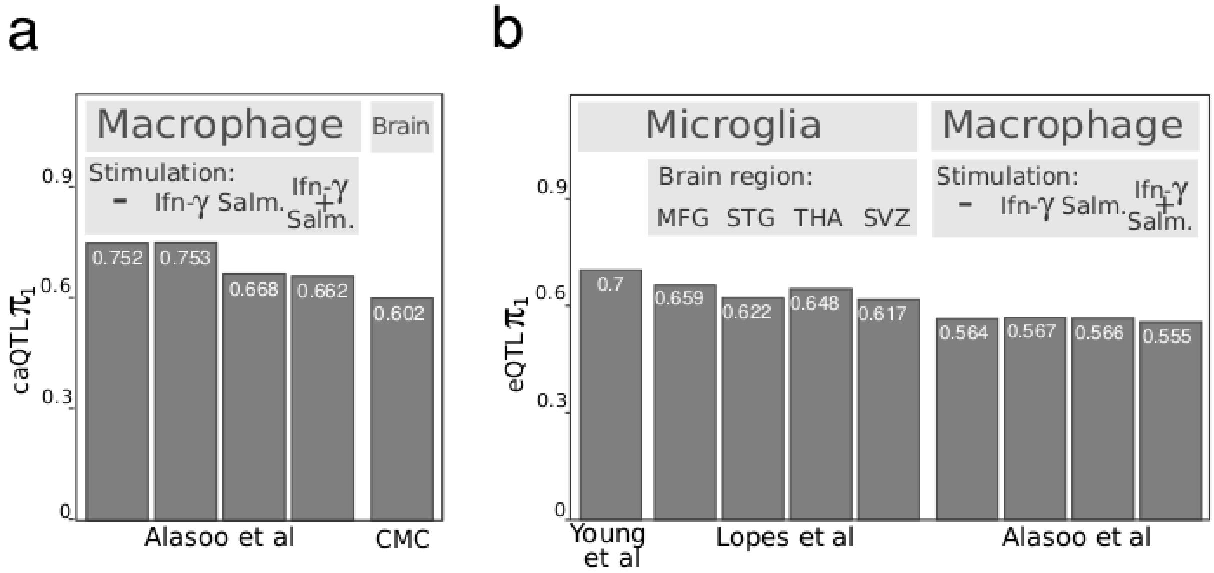


**Fig. S10.**

**Comparison of caQTLs and eQTLs identified in our human microglia to relevant publicly available datasets. A)** $\pi_1$ replication of caQTLs identified in our microglia data to macrophage caQTLs from Alasoo *et al*. [16] and brain homogenate from CommonMind [17]; **B)** $\pi 1$ replication of eQTLs identified in our microglia data to microglia eQTLs from Young *et al*. [7] and Lopes *et al*. [8], and to macrophage eQTLs from Alasoo et.al [16]**.**

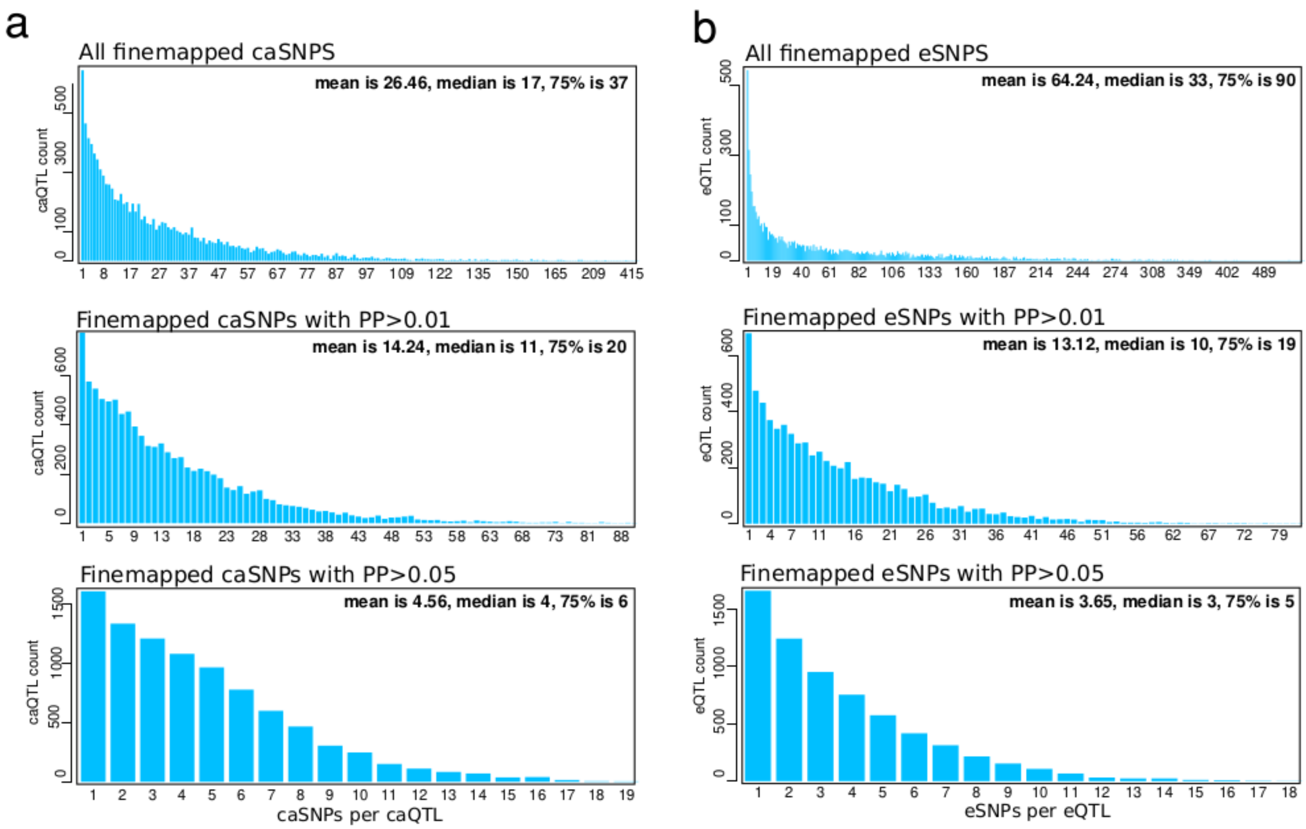


**Fig. S11.**

**Distribution of genetic variants finemapped in caQTL and eQTL analyses.** Distribution of the genetic variants in meta-caQTL and meta-eQTL 95% confidence intervals. Number of **a**) caSNPs and **b**) eSNPs are plotted for all fine-mapped SNPs in significant caQTLs and eQTLs (upper plots: n=269,528/466,348), fine-mapped SNPs with PP >0.01 (middle plots: n=144,592/95,005), and PP >0.05 (lower plots: n=41,403/23,934).

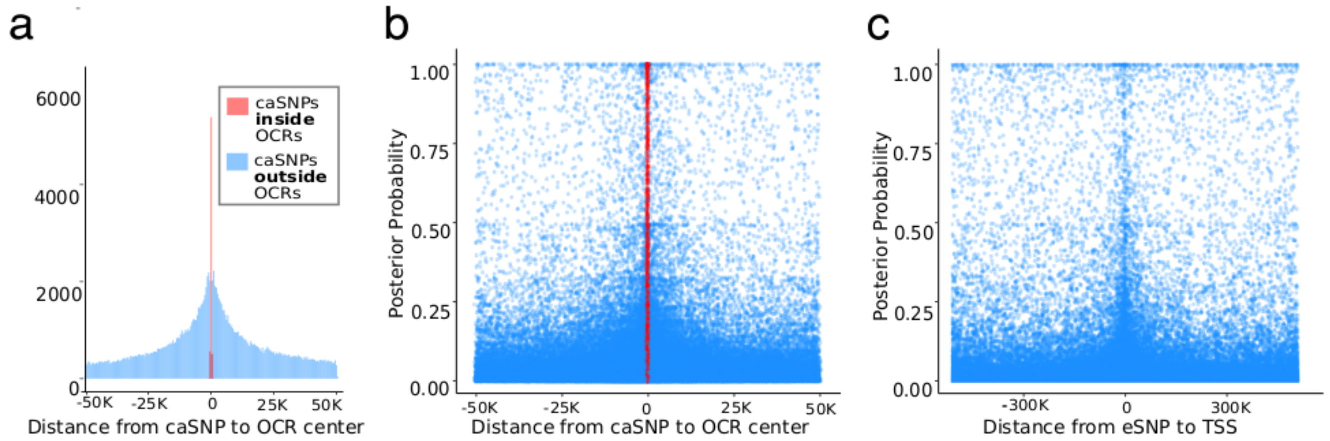


**Fig. S12.**

**Distribution of the caSNPs and eSNPs in relation to the regulated OCRs or genes. a**) Histogram of distances between the OCR centers and caSNPs with PP>0.01 (n=144,592). Red bars indicate caSNPs (n=9,874) located within OCR boundaries (number of unique OCRs = 4,324). **b**) Distance between caSNPs and OCR center vs. caSNPs posterior probability. Red points indicate caSNPs located within the OCR boundaries. **c**) Distance between eSNPs and genes' TSS vs. eSNPs posterior probability.

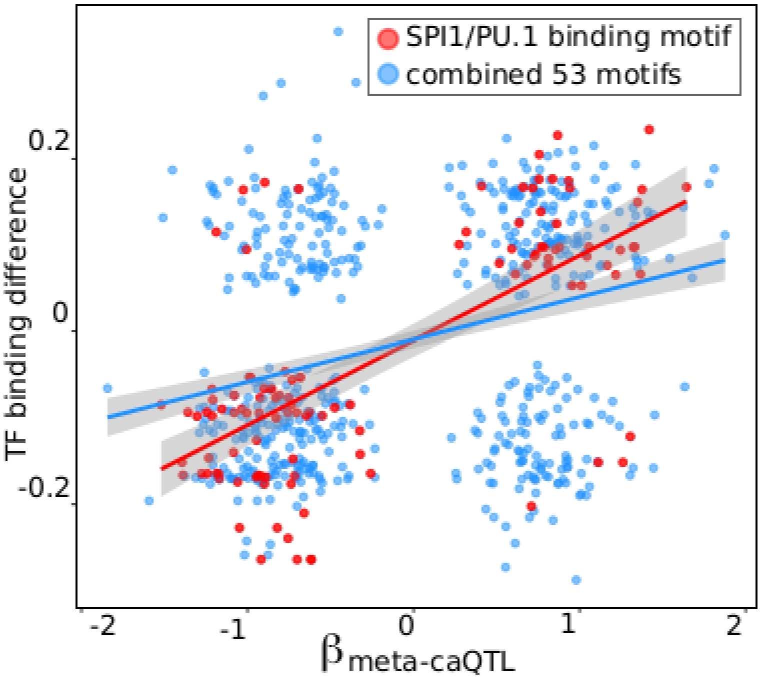


**Fig. S13.**

**Functional impact of caSNPs on TF motif binding ability**. The predicted disruption of TF binding motifs by caSNPs (PP>0.25) as estimated by motifbreakR plotted in relation to the caQTL B coefficient associated with the tested alleles for the 53 TF motifs predicted to be significantly disrupted by caSNPs. The scatter plot includes a total of 675 caSNPs located within OCR boundaries, of which 112 (red points) are located within PU.1 (encoded by SPI1) binding motifs. Relationship between the predicted allelic effect on the binding affinity and caQTL beta coefficient is plotted for caSNPs located within all dysregulated 53 TF motifs (**blue**, $\rho$=0.311, p=1.12x10$^{-16}$, Spearman test) or located within PU.1 motifs (**red**, $\rho$=0.68, p=1.25x10$^{-16}$, Spearman test). Concordance between the predicted disruption of TF binding and negative caQTL coefficient per allele is 0.61 for caSNPs in all TF binding motifs and 0.92 for PU.1 binding motifs.

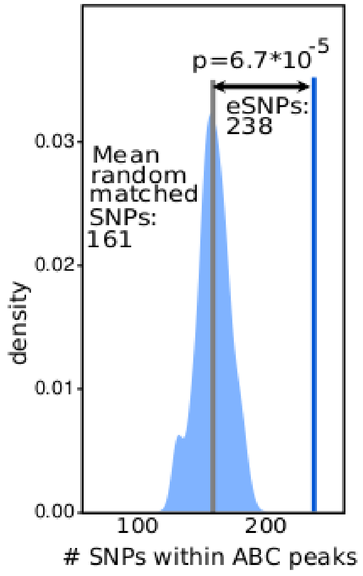


**Fig. S14.**

**OCRs with high ABC scores are enriched for caSNPs compared to SNPs not involved in OCR regulation**. Considering the location of the genetic variants contributing to transcriptional regulation in relation to functional OCRs, we evaluated whether eSNPs are more likely to be located within high ABC OCR peaks compared to genetic variants not linked to a genes' transcriptional regulation. For each of the 7,302 meta e-QTL we selected the maximum of ten eSNPs with the highest posterior probabilities, and identified a non-eQTL SNP at a comparable distance, but in the opposite direction, from the TSS of the e-gene. We observed that 238 fine-mapped eSNPs were localized within $OCR_{ABC}$, 1.48 times higher than a mean of 161 from the same number of matched non-eQTL associated genetic variants ($p = 6.7x10^{-5}$) .

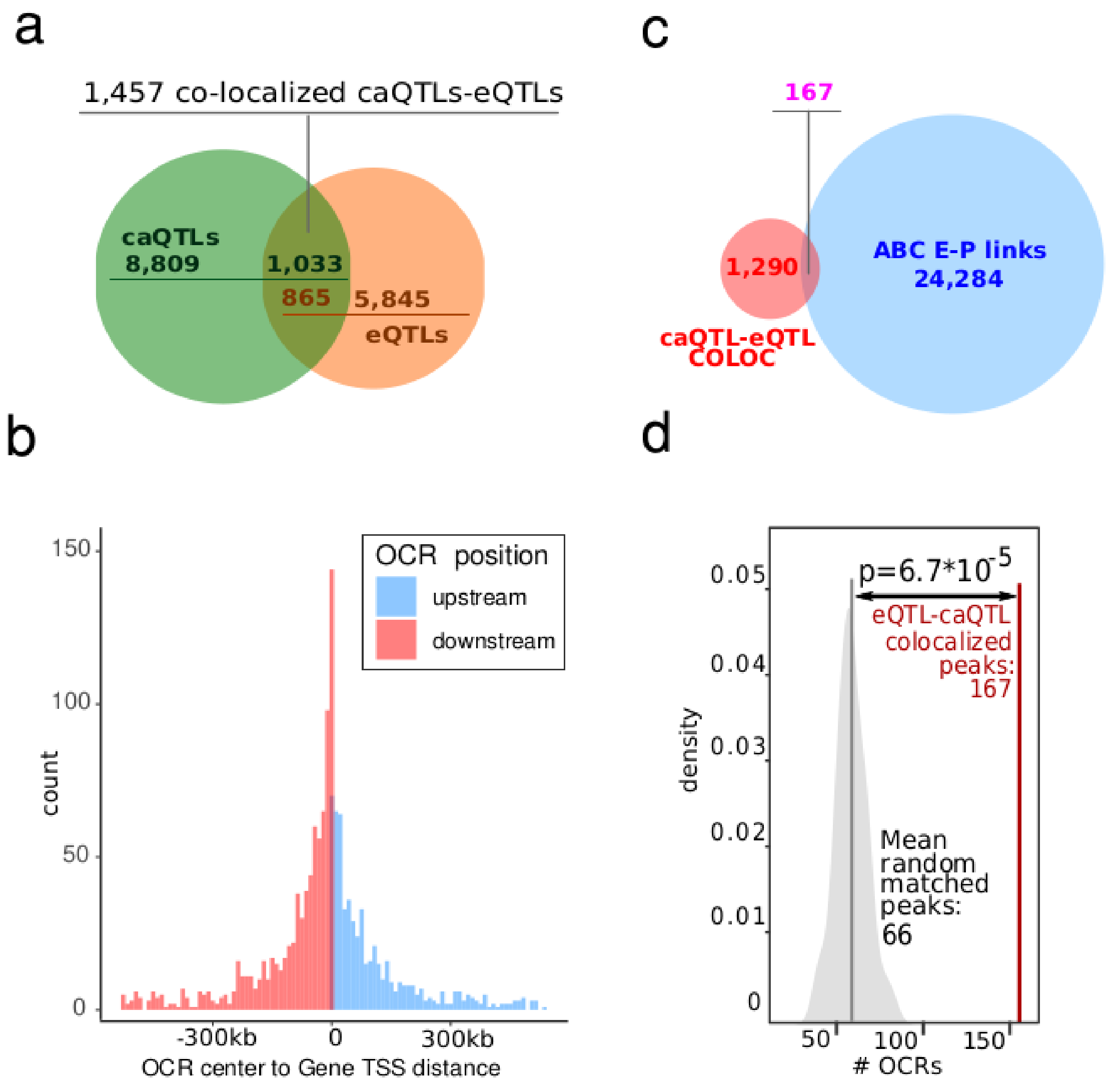


**Fig. S15.**

**The distribution of colocalized eQTLs and caQTLs**. **a)** Venn diagrams represent the colocalized 1,457 OCR-Gene pairs among meta-caQTLs (blue) and meta-eQTLs (red). Colocalized OCR-Gene pairs include 16 OCRs and 14 genes not significant in standalone meta-caQTLs and meta-eQTLs; **b)** The distribution of the distance between the OCRs and genes for the colocalized caQTLs and eQTL pairs. Mean distance is 101kb, and median distance is 53kb. **c)** Overlap between OCR-gene links identified via caQTL-eQTL colocalization method versus E-P links identified via the ABC approach; **d)** CaSNPs for the caQTLs colocalized with eQTLs are more likely to be located within $OCR_{ABC}$ than in random OCRs. Red line represents the number of $OCR_{ABC}$ regulated by colocalized caQTL, versus the distribution of $OCR_{ABC}$ from 100 random sets of 1,457 distance-matched eQTL/caQTL pairs.

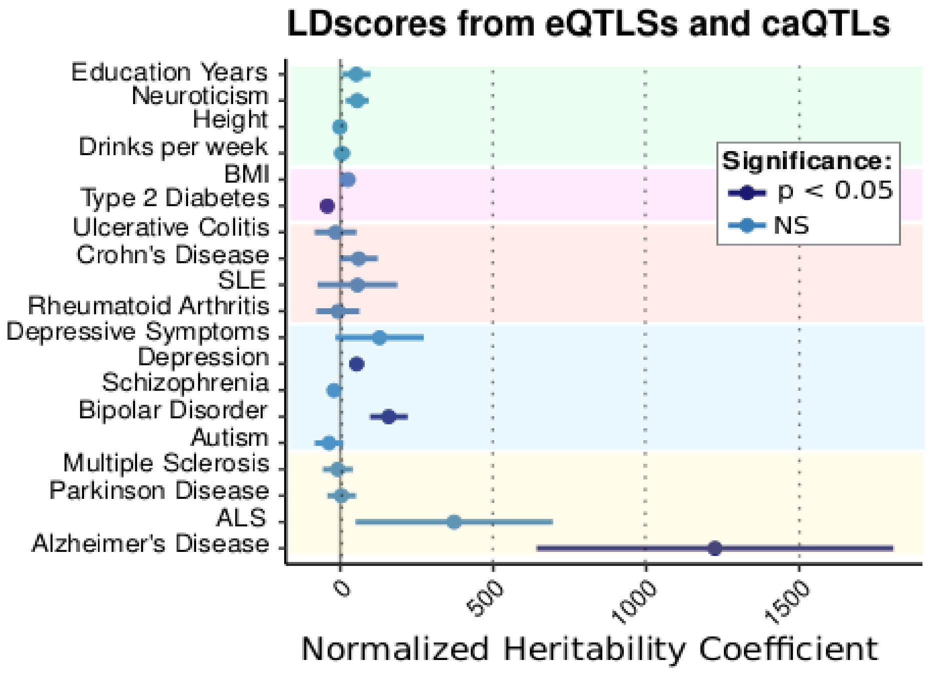


**Fig. S16.**

**LDscore analyses of colocalized caQTL-eQTLs.** Enrichment of SNPs representing 95% confidence interval for the caQTL sets which were colocalized with eQTLs identified in fresh microglia via LDscore analyses using summary statistics from a set of selected GWAS studies representing general, metabolic, inflammatory, psychiatric and neurodegenerative traits. Values in **dark blue** represent nominally significant enrichment. Error bars indicate standard errors for the normalized heritability coefficient.

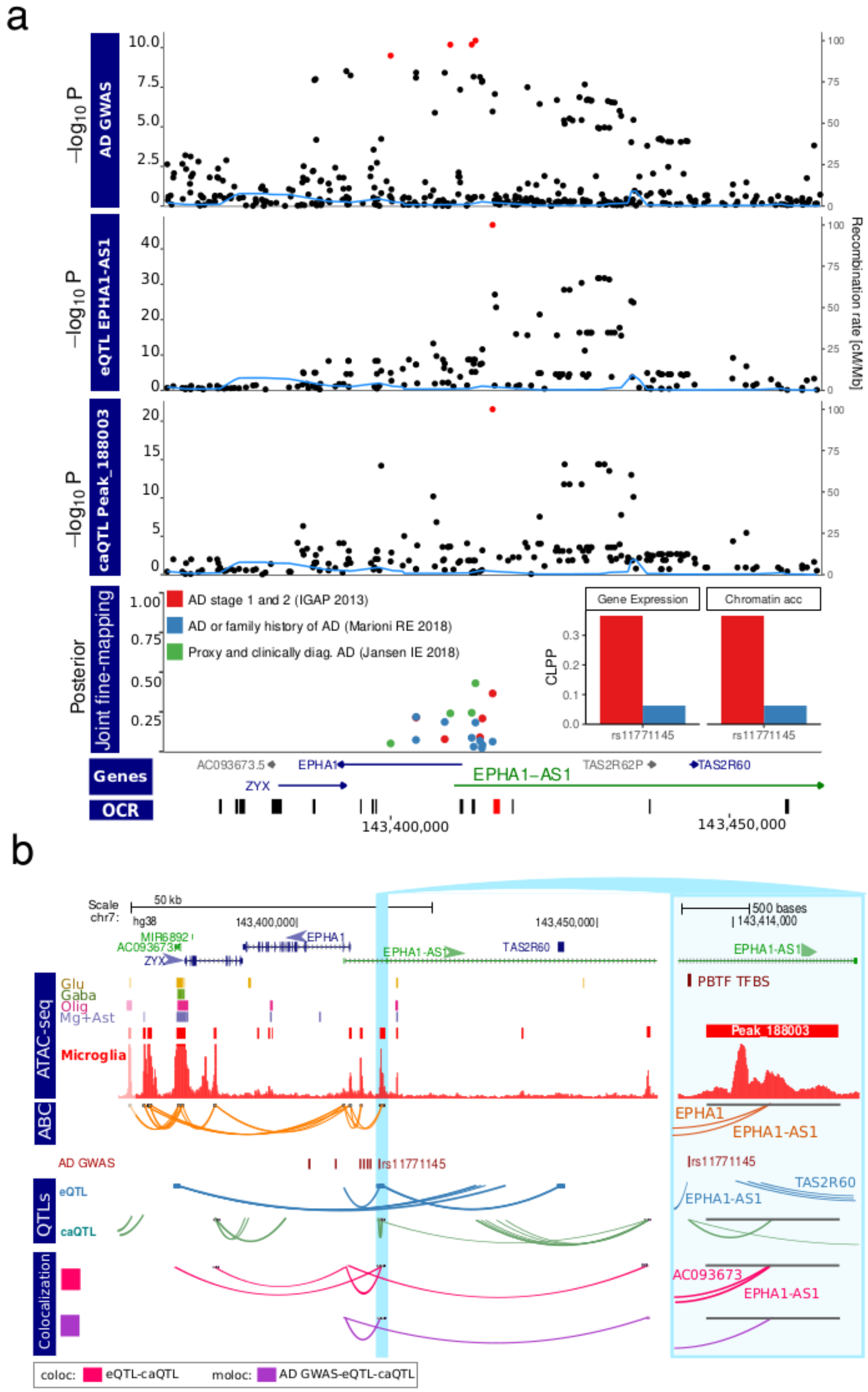


**Fig. S17.**

**Integration of the landscape of AD etiology with genetic regulation of transcriptional and chromatin accessibility in microglia within the *EPHA1-AS1* locus. A)** Local plot showing results from AD GWAS [12], eQTL analysis of *EPHA1/EPHA1-AS1*, and caQTL analysis of peak_188003. CLPP is the joint posterior probabilities (37) between the eSNP/caSNP PP and Jansen AD GWAS PP. Red points indicate genetic variants in the 95% credible set from statistical fine-mapping of each trait. Inset shows colocalization posterior probabilities (CLPP) for the top variant in the credible set for gene expression and chromatin accessibility. **B)** Visualization of *EPHA1/EPHA1-AS1* locus showing: open chromatin regions from 4 cell populations [5] and microglia from this study; E-P interactions (ABC); fine-mapped variants from AD (Jansen) [12]; genetic regulation from eQTLs and caQTL form thus study; and colocalization analysis between pairs of traits (i.e. AD GWAS, gene expression chromatin accessibility) using 'coloc' and all three traits using 'moloc' methods with Jansen *et al*. AD GWAS (pp15>0.5) [27].

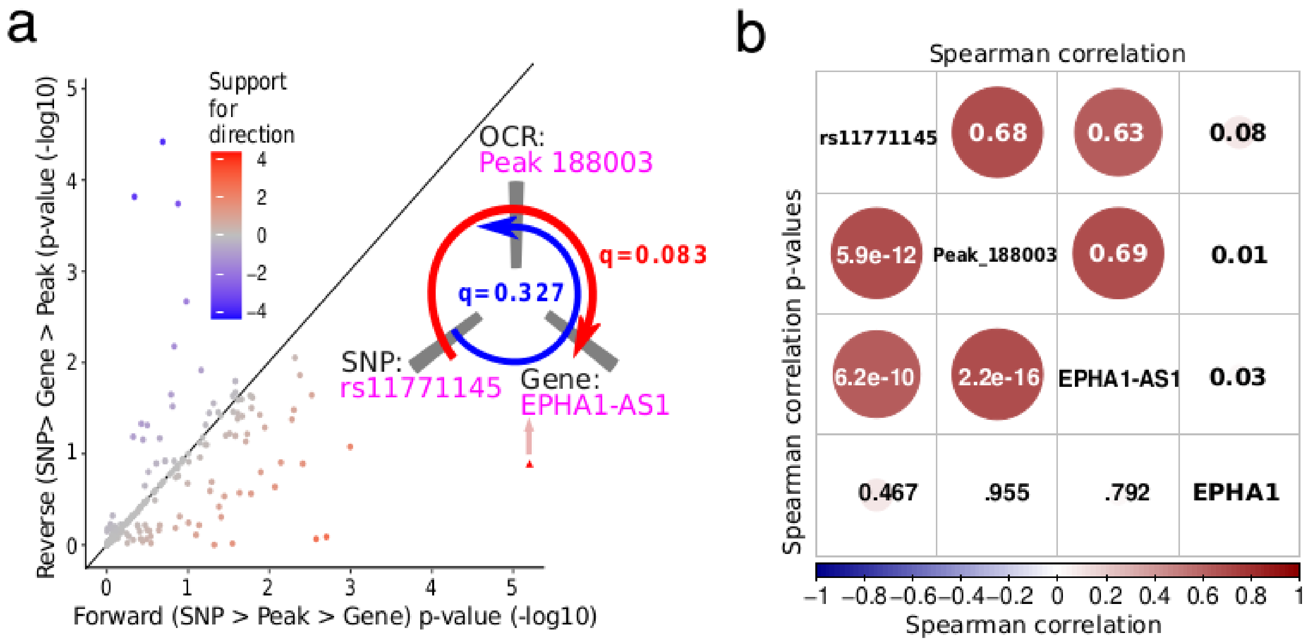


940 **Fig. S18.**

941 **Relationship between colocalized components at EPHA1-AS1 locus.** Relationship between *EPHA1-AS1* expression, OCR peak_188003 and rs11771145. **A)** Application of Causal Inference Test identifies a genetic variant's regulation on transcriptional activity of AD-implicated genes mediated by its effect on chromatin accessibility. **B)** Correlation between the expression of *EPHA1* and *EPHA1-AS1*, ATAC-seq signal at OCR peak_188003 and the genotype of rs11771145.

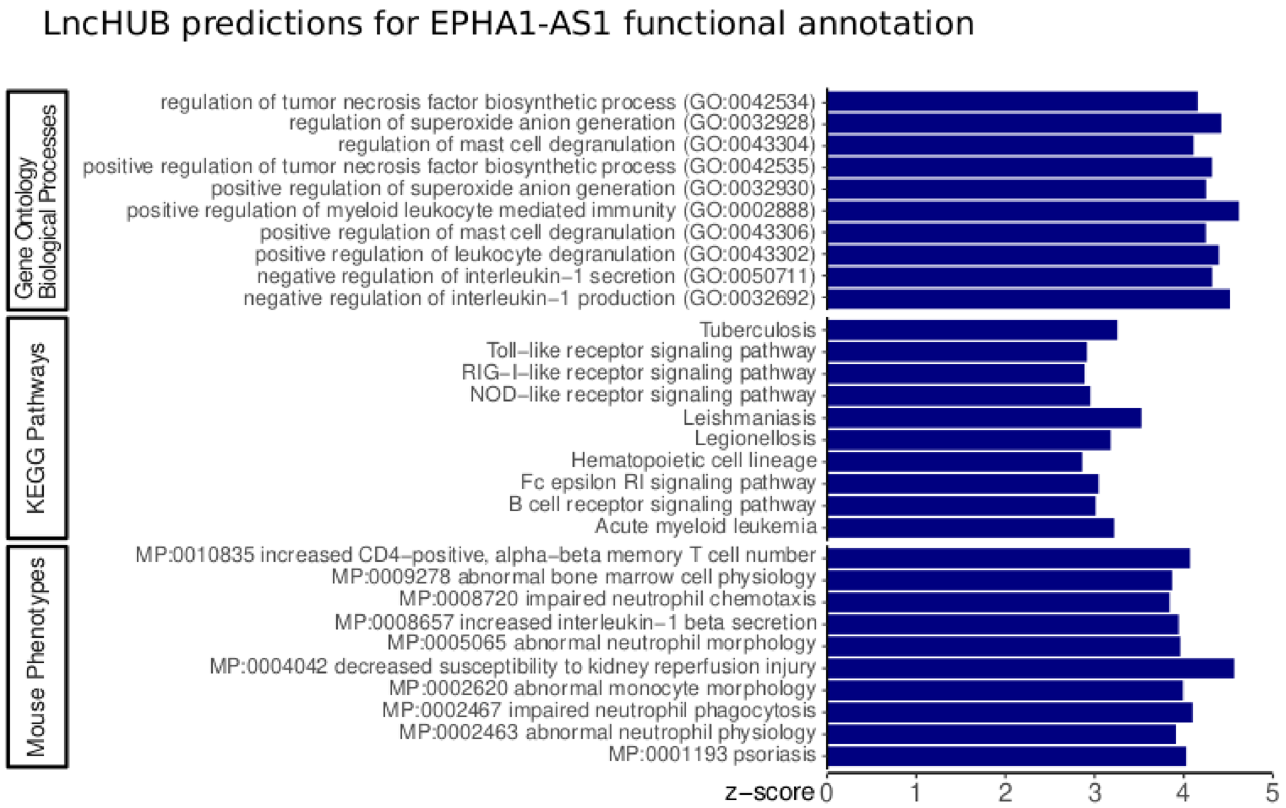


**Fig. S19.**

**Predicted function of EPHA1-AS1 based on coexpression structure.** lncHUB (*44*) predicts gene set annotations of every gene based on genome-wide co-expression structure and known annotations**.** Results are from query https://maayanlab.cloud/lnchub/?lnc=EPHA1-AS1

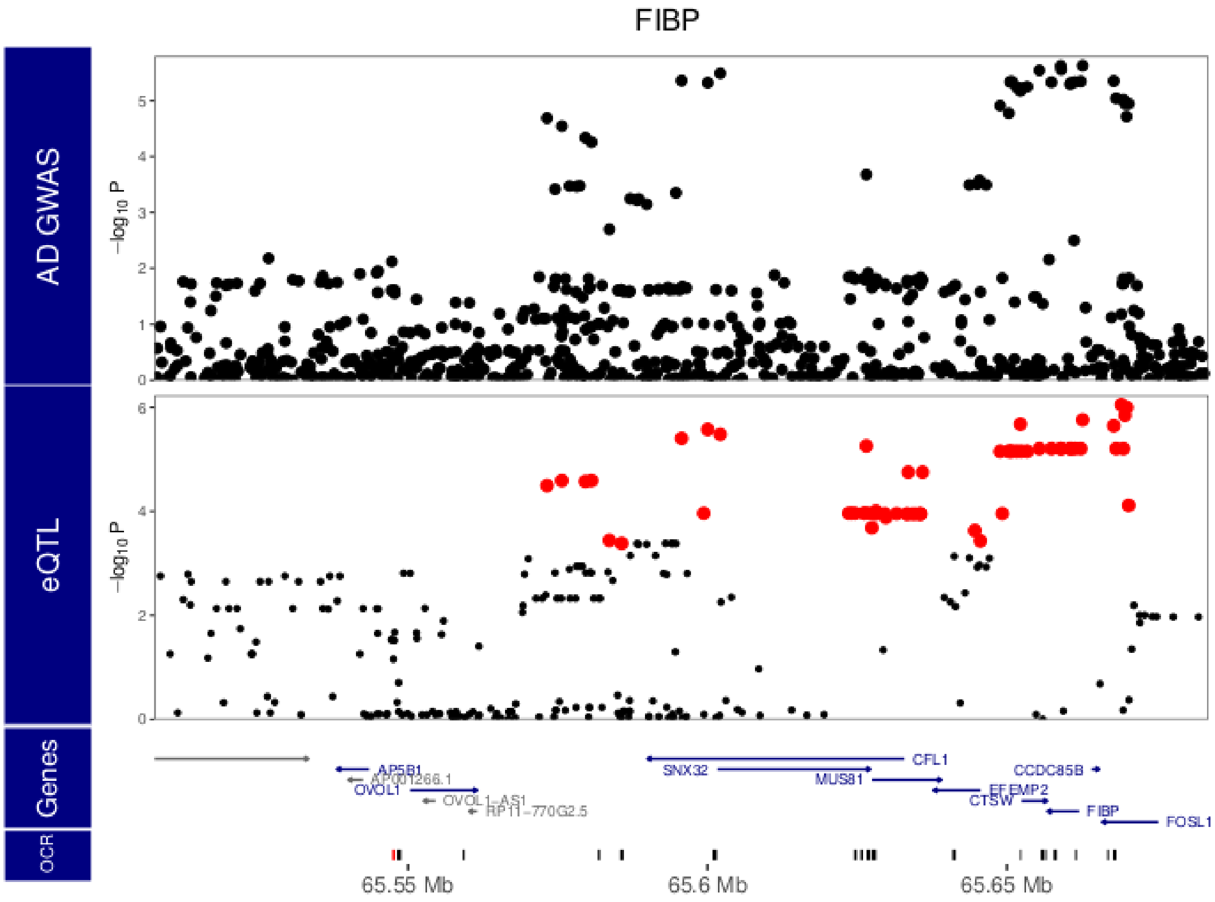


**Fig. S20.**

**Local manhattan plot of AD GWAS [12], and eQTL analysis for *FIBP*. Red** points indicate variants within the 95% confidence interval from statistical fine-mapping.

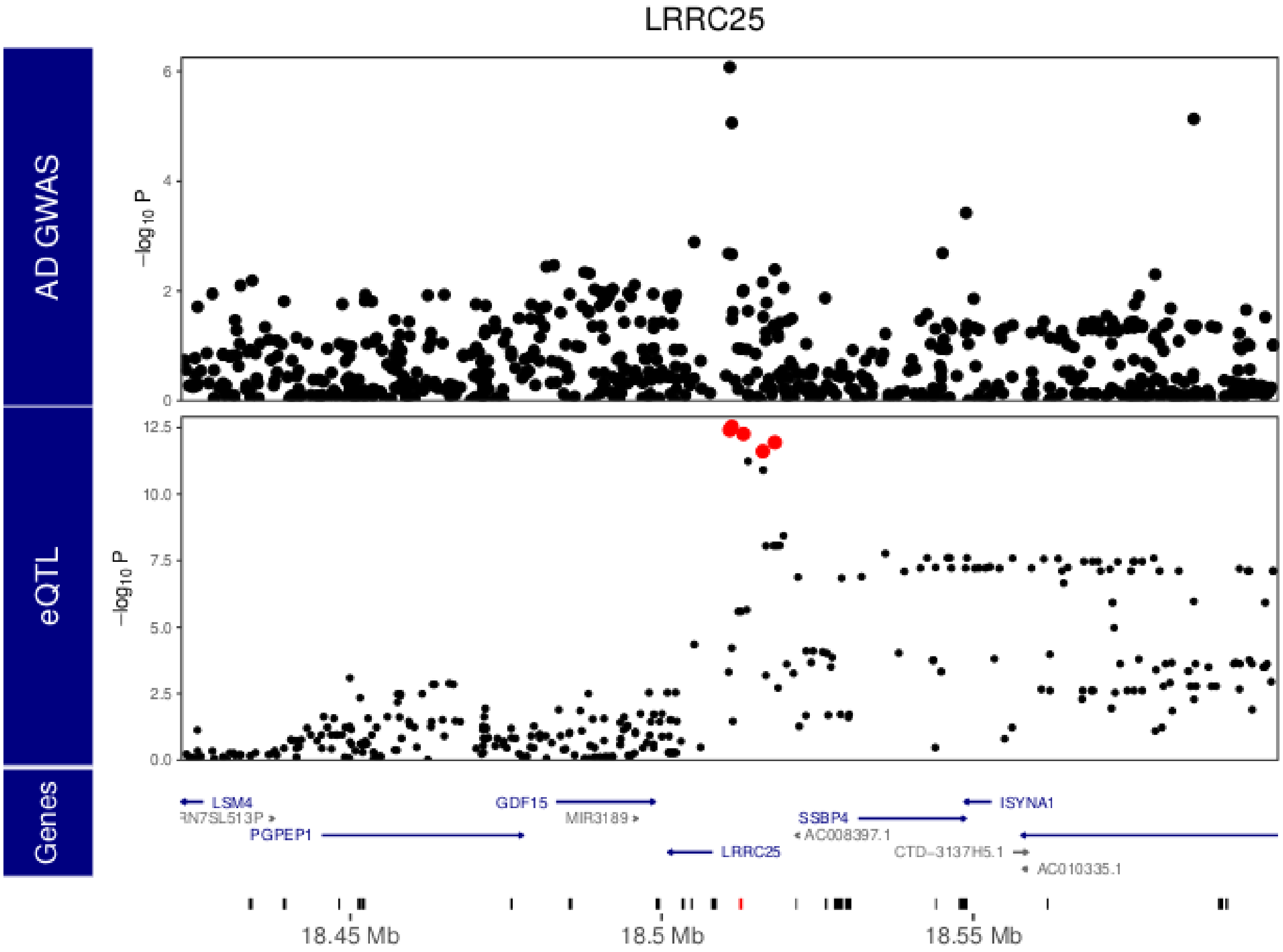


**Fig. S21.**

**Local manhattan plot of AD GWAS** [12]**, and eQTL analysis for *LRRC25*.** Red points indicate variants within the 95% confidence interval from statistical fine-mapping.

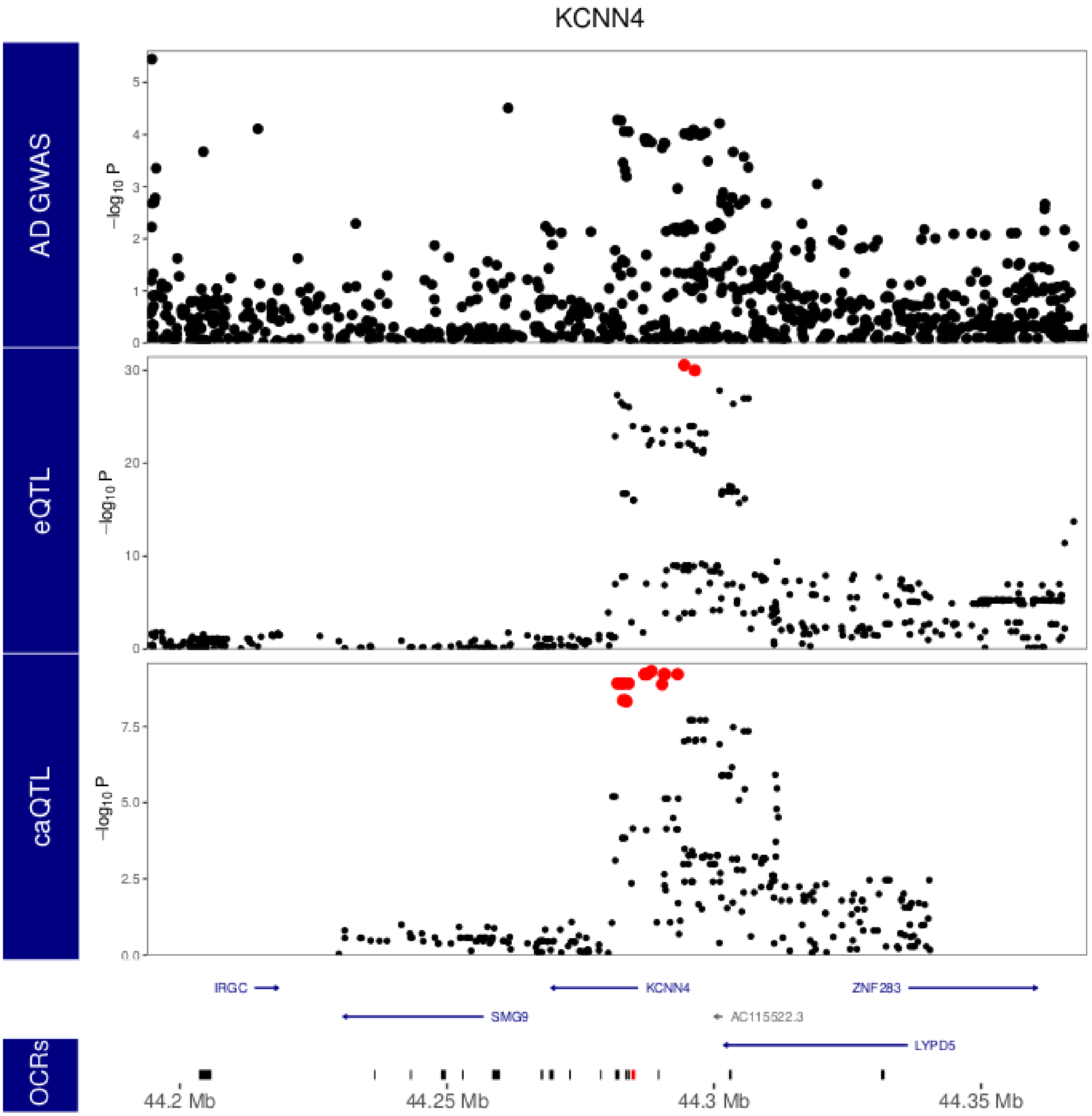


972 **Fig. S22.**

973 **Local manhattan plot of AD GWAS [12,] eQTL analysis for *KCNN4* and caQTL for Peak_82668.**
974 Bottom row indicates open chromatin regions in the window, and the red region indicates the target
975 peak for caQTL analysis shown. **Red** points indicate variants within the 95% confidence interval from
976 statistical fine-mapping.

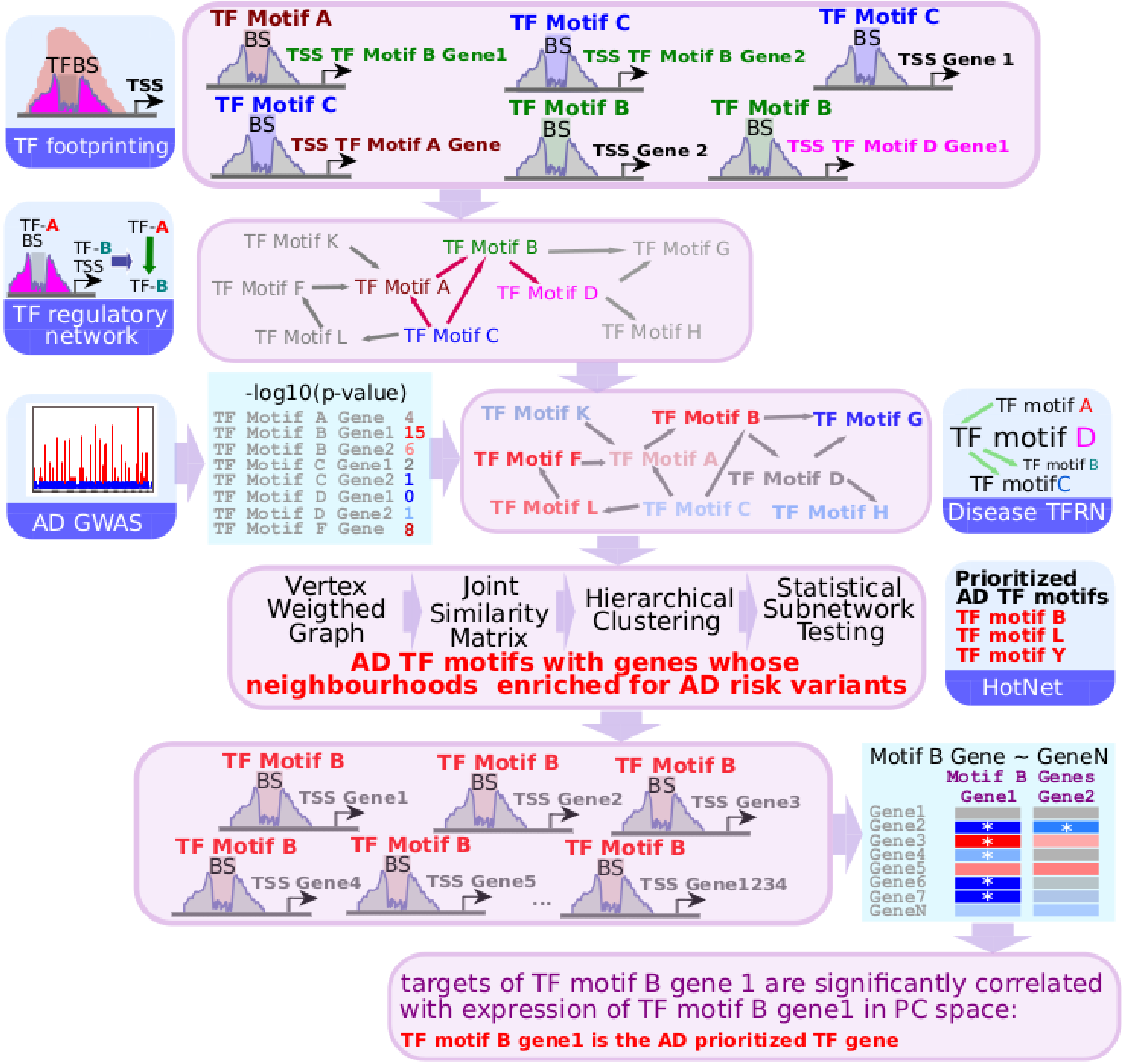


**Fig. S23.**

**Schema of transcription factor binding based analyses to identify AD prioritized TFs.** Bound transcription factors were identified via footprinting analyses [23], and we generated a directed TF regulatory network (TFRN) based on bound TF motifs in the promoter regions of TFs. AD GWAS derived weights [12] were utilised by HotNet [36] to create AD TFRN, within which we identified a subnetwork consisting of 11 TF motifs jointly representing perturbed regulatory hubs in AD. We utilised the available RNAseq data to identify which of the TFs within implicated AD prioritised TF motifs has a significant impact on the downregulated genes, identifying individual TFs relevant to AD biology in the microglia.

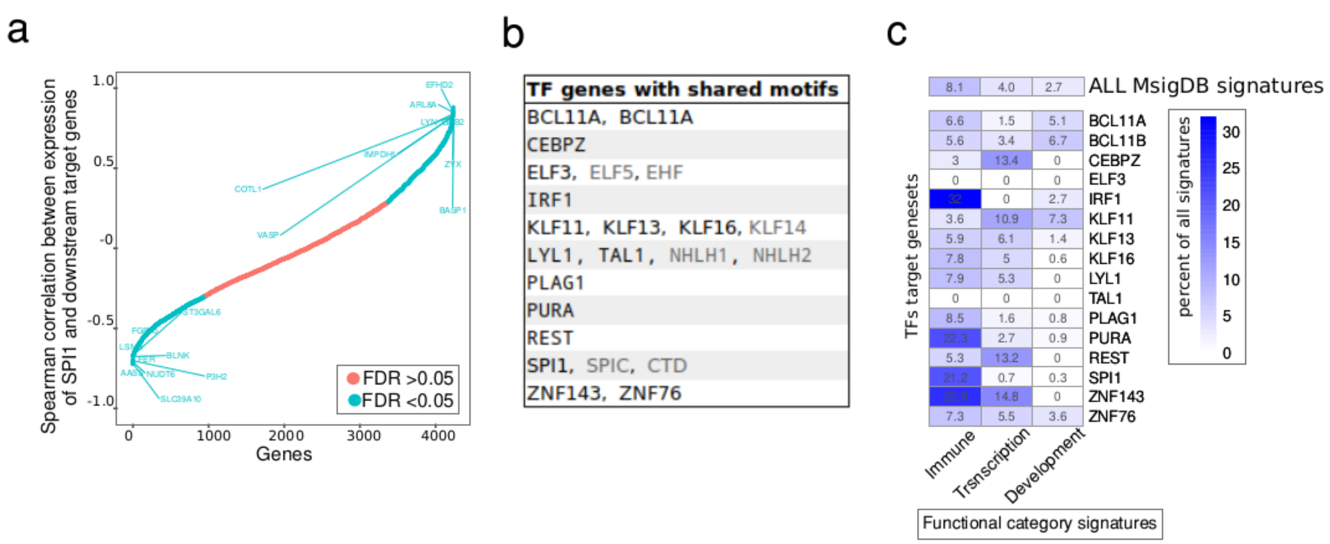


**Fig. S24.**

**Prioritization and functional annotation of individual TF genes among AD prioritised TF motifs.** **A)** Correlation of expression of SPI1 target genes (i.e. 4,226 genes with bound SPI1 TF in their proximal regulatory region) with expression of SPI1 gene itself. Significant correlations are denoted by blue and the names of top ten correlated and the anti-correlated genes are highlighted. **B)** Individual TFs represented by the 11 AD prioritised TF motifs. 16 out of 23 TFs expressed in our microglia data are indicated in black, with grey indicating TF genes below the inclusion threshold (CPM<1 in 20% of all samples). **C)** Functional annotation of the 16 individual TFs from 11 AD prioritized TF motifs by enrichment analyses of the downstream target genes significantly associated (FDR<0.05) with the expression of each TF. Starting from a subset of 8,743 MsigDB signatures, we generated curated subsets including immune-related signatures (curated 755 signatures), transcription-related signatures (curated 245 signatures), and development-related signatures (curated 328 signatures). The values represent the percent of the signature subsets significantly enriched per TF target geneset (FDR<0.05) belonging to each functional subset as a fraction of all significantly enriched signatures.

**Supplementary tables (attached as separate files):**

**Data S1.**

**Sample and patient description.** Demographic and clinical description of donor patients (“Donors_Info”), and relevant technical description of samples utilized in ATAC-seq (“ATAC_QC”), RNA-seq (“RNA_QC”), and Hi-C (“HIC_QC”) analyses.

**Data S2.**

**Description of ATAC-seq datasets utilised in MDS comparisons** (**Fig. 1b**).

**Data S3.**

**Description of the GWAS studies whose summary statistics were utilized in LDscore analyses (Fig. 1f, Fig. 2f, and Fig. 3f)**.

**Data S4.**

**Results of the motifbreakR analysis for the TFs whose binding sites were significantly disrupted by finemapped caSNP**. “Observed” values represent how many caSNPs were predicted to significantly impact the TFs binding affinity by the caSNP, whereas “Random” values represent the number of random genetic variants located within OCRs identified in microglia predicted to significantly impact the TFs binding affinity.

**Data S5.**

**Location distribution of AD snps within microglia OCRs.**

Distribution of AD risk loci fine-mapped in Jansen et. al GWAS with different subsets of microglia OCRs according to their OCR status and distance to genes’ TSS. The considered genetic variants included all SNPs within 95% credible interval, the top 10 or top 5 SNPs per locus, or only SNPs with PP > 0.01 or 0.05.

**Data S6.**

**Summary of colocalization and ABC based analyses for AD GWAS**. “COMBINED_byLocus” includes integrated table aggregating the various analyses per separate AD genomic locus. The other tables include results from each individual analyses utilized in **Fig. 4b**, sorted by the locus chromosomal position.

**Data S7.**

**Distribution of bound TFs in TOBIAS analyses in Microglia, GABAergic and Glutamatergic Neurons, and Oligodendrocytes**.

**References utilized in supplementary data only**:
